# Integrating PCR-free amplification and synergistic sensing for ultrasensitive and rapid CRISPR/Cas12a-based SARS-CoV-2 antigen detection

**DOI:** 10.1101/2021.06.17.21258275

**Authors:** Xiangxiang Zhao, Zhengduo Wang, Bowen Yang, Zilong Li, Yaojun Tong, Yuhai Bi, Zhenghong Li, Xuekui Xia, Xiangyin Chen, Weishan Wang, Gao-Yi Tan, Lixin Zhang

## Abstract

Antigen detection provides particularly valuable information for medical diagnoses; however, the current detection methods are less sensitive and accurate than nucleic acid analysis. The combination of CRISPR/Cas12a and aptamers provides a new detection paradigm, but sensitive sensing and stable amplification in antigen detection remain challenging. Here, we present a PCR-free multiple trigger dsDNA tandem-based signal amplification strategy and a de novo designed dual aptamer synergistic sensing strategy. Integration of these two strategies endowed the CRISPR/Cas12a and aptamer-based method with ultra-sensitive, fast, and stable antigen detection. In a demonstration of this method, the limit of detection was at the single virus level (0.17 fM, approximately two copies/μL) in SARS-CoV-2 antigen nucleocapsid protein analysis of saliva or serum samples. The entire procedure required only 20 minutes. Given our system’s simplicity and modular setup, we believe that it could be adapted reasonably easily for general applications in CRISPR/Cas12a-aptamer-based detection.

## 1. Introduction

Antigens have been used as diagnostic hallmarks for many diseases, particularly infection with viruses, for example, severe acute respiratory syndrome coronavirus 2 (SARS-CoV-2), the cause of the current global coronavirus disease 2019 (COVID-19) pandemic ^[1]^. Many methods have been developed for SARS-CoV-2 detection through nucleic acids, antigens, and antibodies. Although PCR-based nucleic acid detection is the most accepted and reliable approach, it is relatively slow and device-dependent, owing to the requirements of RNA extraction and reverse transcription. In addition, RNA is less stable than protein (antigen) in laboratory conditions ^[2, 3]^. Moreover, antibody-based tests work well only in late stages of infection, after patients have mounted immune responses, and therefore are not suitable for early diagnosis ^[4]^. In comparison, antigen-based SARS-CoV-2 detection has several unique merits in COVID-19 diagnosis. Given its robustness and simplicity, it provides an important addition to the toolbox for SARS-CoV-2 detection ^[3, 5, 6]^. However, compared with mature nucleic acid detection approaches, recent state-of-the-art methods for SARS-CoV-2 antigen detection have some deficiencies in low abundance viral sample detection (*e*.*g*., early diagnosis) ^[3]^. The underlying reason is that signal amplification of low amounts of antigen is not possible. In contrast, the amount of antibodies are already amplified *in vivo* after antigen stimulation, and nucleic acids can be amplified *in vitro* by PCR or LAMP (loop-mediated isothermal amplification) ^[7]^, thus boosting both signals. To address the challenge of signal amplification for antigen detection, we endeavor to develop a novel antigen detection system with high sensitivity and excellent performance by amplifying the sensing signals.

CRISPR/Cas-based nucleic acid detection systems, such as SARS-CoV-2 DETECTR ^[8]^, SHEROLOCK ^[9]^, STOPCovid ^[10]^, and AIOD-CRISPR ^[11]^, have been developed and applied in rapid diagnostic tests for SARS-CoV-2. The advantages of CRISPR/Cas-based detection systems have been well demonstrated, and these methods have shown great potential during the COVID-19 pandemic, owing to their superior speed, portability, low cost, and comparable sensitivity to that of the traditional gold standard of RT-qPCR assays (one copy/μL of viral RNA) ^[12, 13, 14]^. However, the CRISPR/Cas-based detection paradigm has not been expanded to SARS-CoV-2 antigen detection ^[15]^. To fill this gap, we proposed that the abovementioned antigen detection challenges could be overcome by coupling CRISPR/Cas and a *de novo* designed highly efficient and stable signal amplification system.

Aptamers as a general biosensing platform can be applied to biosensing or detection of a broad range of analytes ^[16]^. Previously, CRISPR/Cas12a and aptamer-mediated systems for detection of diverse analytes (*e*.*g*., proteins or small molecules) have been developed ^[12, 17]^. These platforms directly translate the signal of aptamer recognition target molecules into a CRISPR-mediated nucleic acid detection-based output signal. However, owing to the lack of stable amplification of the input signal, the detection sensitivity this method has been unable to meet the requirements for low abundance sample detection or early diagnosis of COVID-19, in which the viral titer is usually as low as several copies per microliter (*e*.*g*., in saliva) ^[18]^.

Here, to improve the sensitivity or sensing efficiency of the CRISPR/Cas12a and aptamer-mediated method and achieve ultrasensitive detection, we innovated two aspects for the configuration of this new antigen detection platform. First, we developed a PCR-free amplification strategy to convert the aptamer-antigen recognition signal to more trigger dsDNA signal. Second, we undertook *de novo* design of a dual aptamer synergetic module to more sensitively sense the antigen independently of the affinity between the aptamer and antigen. In a demonstration of this platform, we successfully identified two copies/μL SARS-CoV-2 in samples by using our newly established system of antigen detection; the LOD was comparable to that of the gold standard of PCR-based nucleic acid detection, but the detection was more rapid (∼ 20 min) and the costs were lower.

## 2. Results

### 2.1 Design and construction of a prototype CRISPR/Cas12a and aptamer-mediated platform for SARS-CoV-2 antigen detection

The SARS-CoV-2 viral surface contains four types of proteins, *i*.*e*., spike (S), nucleocapsid (N), membrane (M), and envelope (E), which have been used in antigen detection. In this study, to couple antigen recognition with a CRISPR-based detection system, we used a specific aptamer (A48, with an equilibrium dissociation constant (*K*_D_) of 0.49 nM) ^[19]^ of N protein as the recognition element to develop a SARS-CoV-2 antigen biosensor with our recently established CRISPR/Cas12a-based biosensing platform ^[12]^. Our prototype was denoted the antigen biosensing platform CaT-Smelor-Covid.v1 (CRISPR/Cas12a and aptamer-mediated detector of diverse analytes for COVID-19, version 1). The working principle of CaT-Smelor-Covid.v1 is illustrated in Fig. 1a. In brief, a hybrid DNA (HyDNA) containing the Cas12a triggering dsDNA and a single-stranded DNA (ssDNA) complementary to parts of the aptamer A48 sequence was anchored to A48-coated magnetic beads (MB) (Supplementary Fig. 1). SARS-CoV-2 nucleocapsid protein (Np), if present, interacts with A48 and releases the HyDNA. The released HyDNA then triggered the collateral ssDNA cleavage activity of Cas12a and then cut the fluorophore quencher (FQ)-labeled ssDNA probe, thus producing a readable fluorescence signal output. After optimizing the reaction system and signal-to-noise (S/N) ratio (Supplementary Fig. 2, 3), we found that the linear detection range of Np was 0.19–781 pM (*R*^2^ > 0.99) (Fig. 1b, c, Supplementary Fig. 4) with a LOD of 32 fM (approximately 193 copies/μL). For nucleic acid detection, the LOD of the gold-standard RT-qPCR and CRISPR-based methods reached up to one copy/μL of viral RNA ^[6]^. Thus, this prototype did not meet the requirements for detection of SARS-CoV-2 in saliva samples or early diagnosis. Developing novel strategies for sensitive sensing and stable amplification is key for this CRISPR-based antigen detection paradigm.

**Figure 1.**
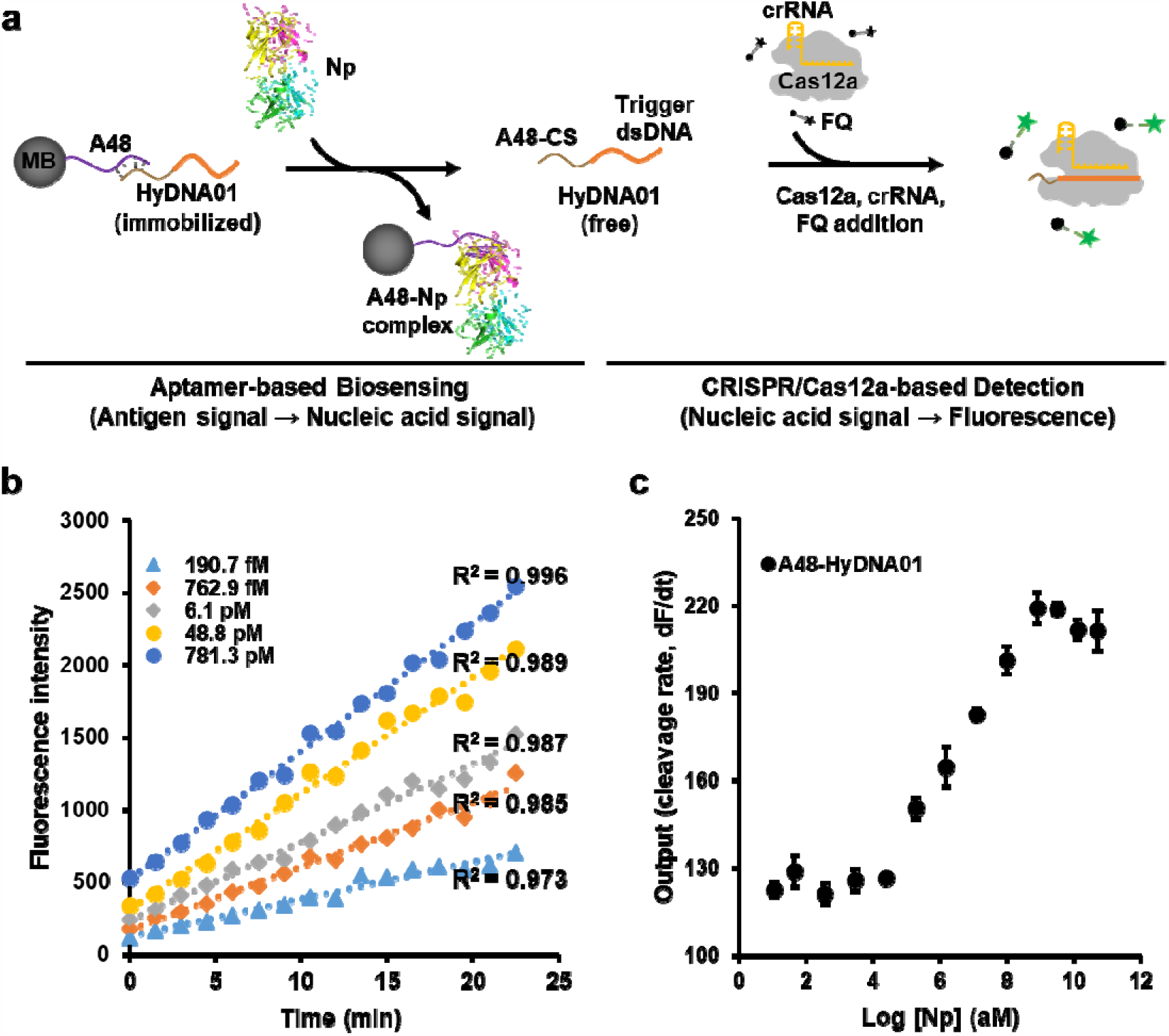
Detection of Np with the CaT-Smelor-Covid.v1 platform. **(a)** Schematic diagram of CaT-Smelor-Covid.v1. MB: streptavidin-coated magnetic bead. A48: aptamer targeting the SARS-CoV-2 nucleocapsid protein. HyDNA01: HyDNA containing a single Cas12a trigger dsDNA. Np: nucleocapsid protein. **(b)** Readout of the fluorescence signal by the CaT-Smelor-Covid.v1 platform in response to Np. The background signal (control) has been subtracted from the values displayed in the graph. **(c)** Relationship between the slope and Np concentration output with the prototype antigen biosensing platform. A48-HyDNA01: core component of the CaT-Smelor-Covid.v1 platform. The data are the means and standard error (SE) of three independent replicates.

### 2.2 Introduction of a PCR-free strategy to amplify the CRISPR/Cas12a generated signal

In the CRSIR/Cas12a and FQ-based signal output and display modules, the slopes of the fluorescence signal are proportional to the concentration of the released triggering dsDNA ^[12]^. To increase or amplify the signal generated by the Cas12a triggering dsDNA, we designed a cluster of multiple triggering dsDNAs in tandem. Considering the trade-off between the S/N ratio and amplified signal, we kept the length of dsDNA below 600 bp, a length accommodating as many as 10 copies of the Cas12a triggering dsDNA with a 30 bp interval sequence (Supplementary Fig. 5). Then the HyDNA containing different copy numbers (one to ten) of Cas12a triggering dsDNAs was anchored to A48-coated MB (Supplementary Fig. 6). The slopes of the fluorescence signal were proportional to one to ten copies of Cas12a triggering dsDNA (HyDNA10; Fig. 2c, d, Supplementary Fig. 7). Thus, we constructed CaT-Smelor-Covid.v2 (version 2) by using HyDNA10 instead of HyDNA01 in the CaT-Smelor-Covid.v1 (Fig. 2a, b). When detecting Np, we observed that the slope of the fluorescence intensity was linear (*R*^2^ > 0.99), and the detection range was 6–381 fM (Fig. 2e, f, Supplementary Fig. 8) with a LOD of 3.48 fM (approximately 42 copies/μL), thus indicating that the introduction of clustered multi-copy Cas12a triggering dsDNA indeed boosted the signal. With the introduction of a PCR-free signal amplification procedure, CaT-Smelor-Covid.v2 showed the expected characteristics of high stability and rapid detection. However, the LOD was inadequate for real-world detection in saliva samples or early diagnosis.

**Figure 2.**
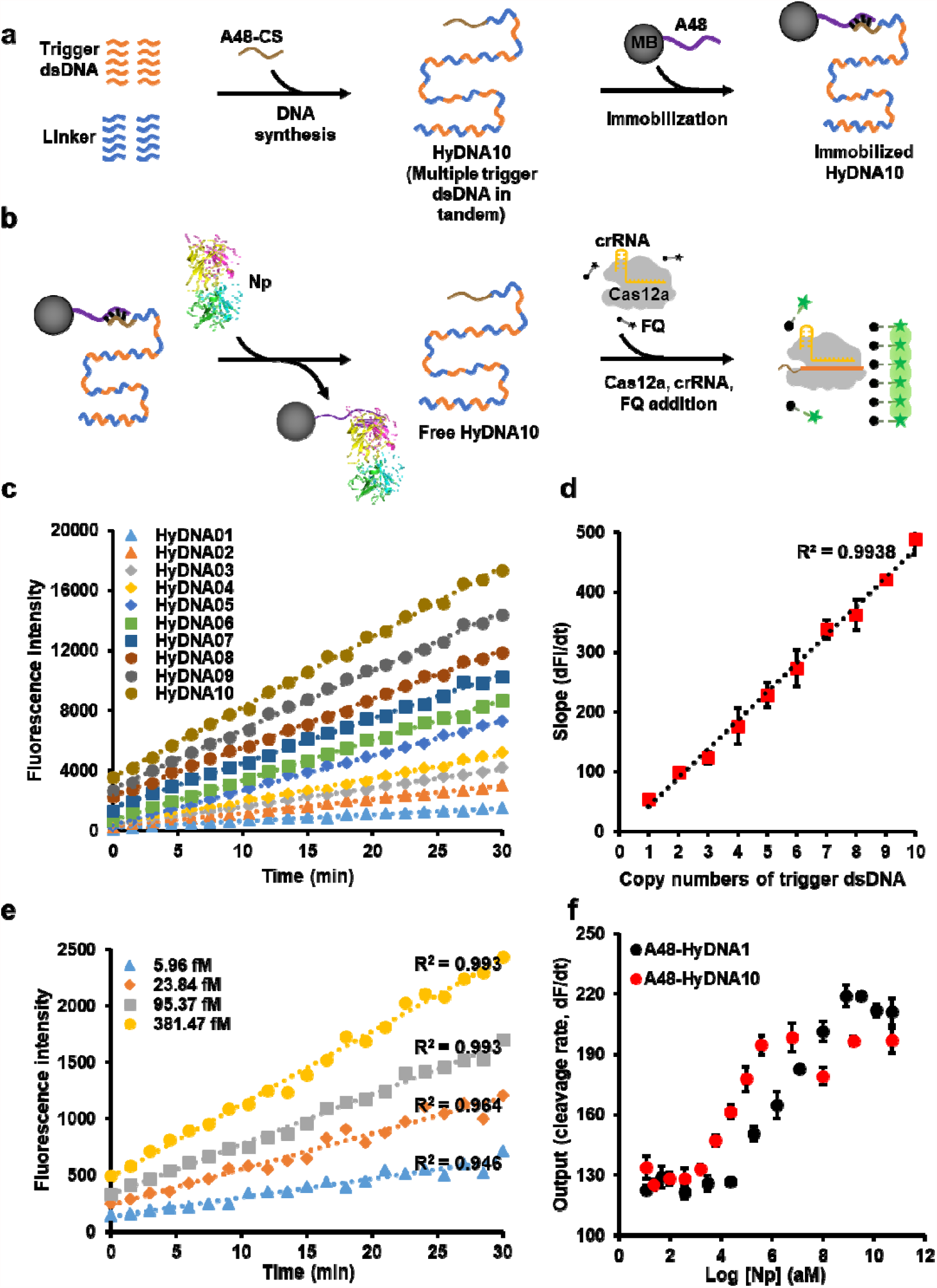
Detection of Np with the CaT-Smelor-Covid.v2 platform. **(a)** Constructing schematic diagram of cluster of multiple triggering dsDNAs in tandem. HyDNA10: HyDNA containing ten Cas12a trigger dsDNAs. **(b)** Sensing schematic diagram of the CaT-Smelor-Covid.v2 platform. **(c)** Plot of the fluorescence level against the HyDNAs. **(d)** The slopes of the fluorescence signal are proportional to the copy numbers of Cas12a trigger dsDNA. **(e)** Readout of the fluorescence signal by the CaT-Smelor-Covid.v2 platform in response to Np. **(f)** Relationship between the slope and Np concentration output with the CaT-Smelor-Covid.v2 platform and the CaT-Smelor-Covid.v1 platform. In c and e, data are the means and SD of three independent replicates.

### 2.3 Introduction of Y-shaped DNA-based aptamers to synergistically increase the sensing sensitivity

To further improve the detection sensitivity of CaT-Smelor-Covid.v2, we pursued a *de novo* synergistic strategy in CaT-Smelor-Covid.v3 (version 3) by applying two different aptamers recognizing different epitopes of Np to release two cognate tandem HyDNA10 molecules (Fig. 3a, b). When one aptamer binds the antigen and releases HyDNA, it promotes the response of the other aptamer to the same antigen and the release HyDNA due to proximity, thus producing a synergistic effect. Here, we introduced a biotin labeled Y-shaped DNA to simultaneously anchor the aptamer (A48) and another previously characterized aptamer (A61, *K*_D_ = 2.74 nM) ^[19]^ on the MB. Then the two optimized HyDNA10 molecules (Supplementary Fig. 9) were anchored on MB via interaction with aptamers A48 and A61. With CaT-Smelor-Covid.v3, we achieved excellent Np detection. Trace amounts of Np at concentrations of 0.19–2.98 fM were unambiguously and readily detected, with an LOD of 0.17 fM (Fig. 3c, d, Supplementary Fig. 10). The LOD of SARS-CoV-2 was approximately two copies/μL, the lowest value reported to date for SARS-CoV-2 antigen detection through a CRISPR/Cas-based method ^[3, 15]^. As expected, the response signal of CaT-Smelor-Covid.v3 allowed us to calculate a Hill coefficient of approximately 1.5, thus indicating positive cooperativity.

**Figure 3.**
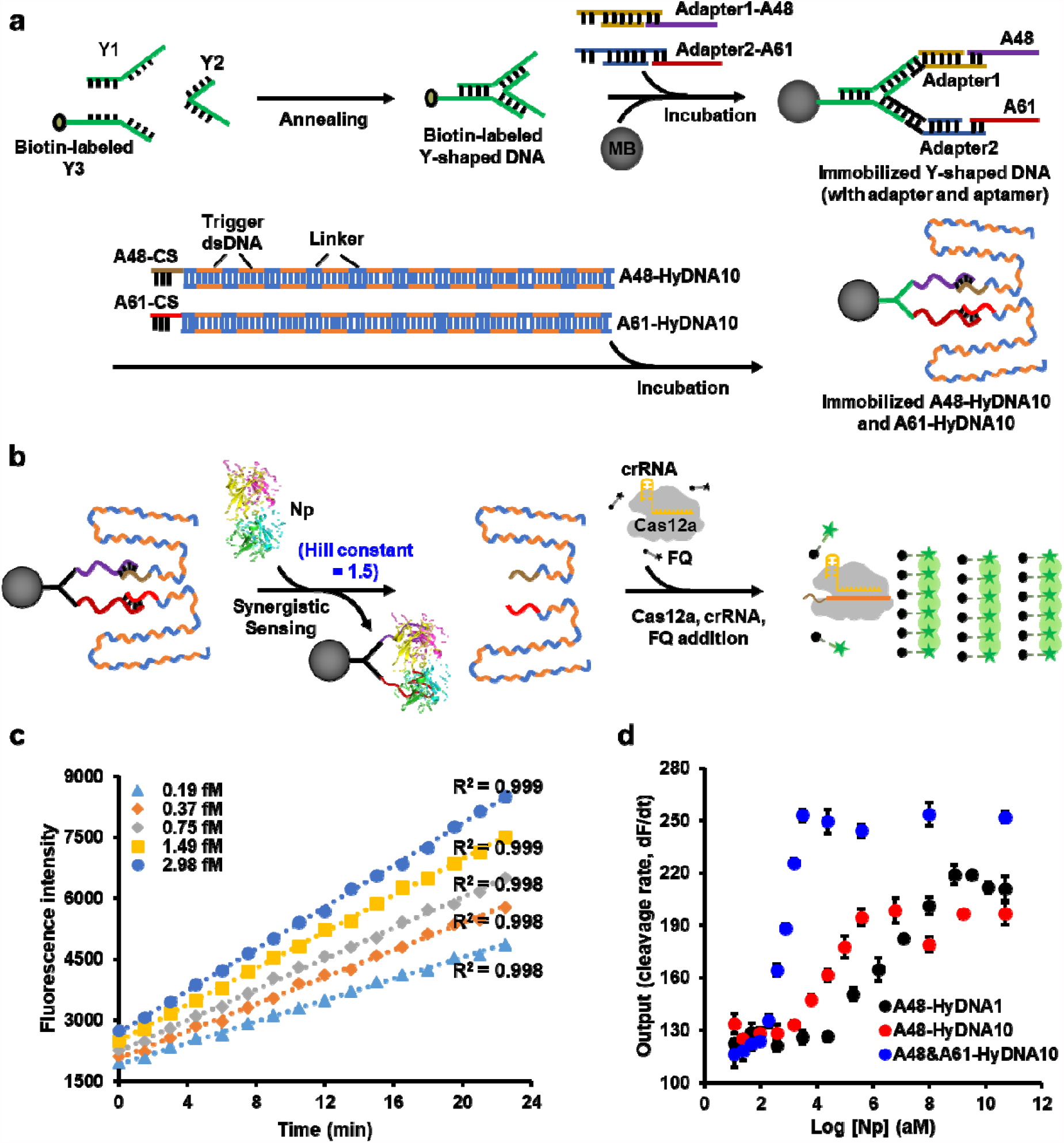
Detection of Np with the CaT-Smelor-Covid.v3 platform. **(a)** Design and construction of the dual aptamer synergetic module. Y1, Y2 and biotin labeled Y3 oligonucleotides were complementary to one another. A mixture of the same molar amount of these oligonucleotide strands formed a biotin-labeled Y-shaped DNA. The MB-Y-A48+A61 complex was constructed through mixture of Y-DNA, MB, Adapter1-A48, and Adapter2-A61. Assembly of the Y-A48+A61-HyDNA10 complex was accomplished through mixture of MB-Y-A48+A61, com10-HyDNA10, and com11-HyDNA10. **(b)** Sensing schematic diagram of the CaT-Smelor-Covid.v3 platform. **(c)** Readout of the fluorescence signal by the CaT-Smelor-Covid.v3 platform in response to Np. **(d)** Relationship between the slope and Np concentration output of the biosensing platforms. The data are the means and SD of three independent replicates.

### 2.4 Rapid and sensitive detection of inactivated SARS-CoV-2 in clinical samples

The LOD of CaT-Smelor-Covid.v3 for antigen detection was very close to that of the gold standard of RT-qPCR and CRISPR/Cas-based nucleic acid detection ^[8, 10]^, thus indicating that this platform had great potential for real-life applications. We then tested the performance of our CaT-Smelor-Covid.v3 on authentic sample detection by using inactivated SARS-CoV-2 ^[20]^. As shown in Fig. 4a, as few as two copies/μL SARS-CoV-2 could be efficiently detected (*P* < 0.05). Moreover, the entire process required only approximately 20 minutes with a commercially available portable fluorescence detector (Qubit 4 Fluorometer, Thermo Fisher Scientific, US) (Fig. 5). To demonstrate the performance of our method, we spiked saliva and human serum samples with different concentrations of inactivated SARS-CoV-2 for Np detection. In one single-blind test, 30 virus-spiked and 30 virus-unspiked saliva samples were detected with a Qubit 4 Fluorometer, and all positive samples were detected (*P* < 0.001; Fig. 4b, Supplementary Fig. 11–13). In another single-blind test, our method correctly distinguished the 30 positive and 30 negative samples in spiked human serum (*P* < 0.001; Fig. 4b). These results indicated that CaT-Smelor-COVID.v3 was ultrasensitive and stable for SARS-CoV-2 detection.

**Figure 4.**
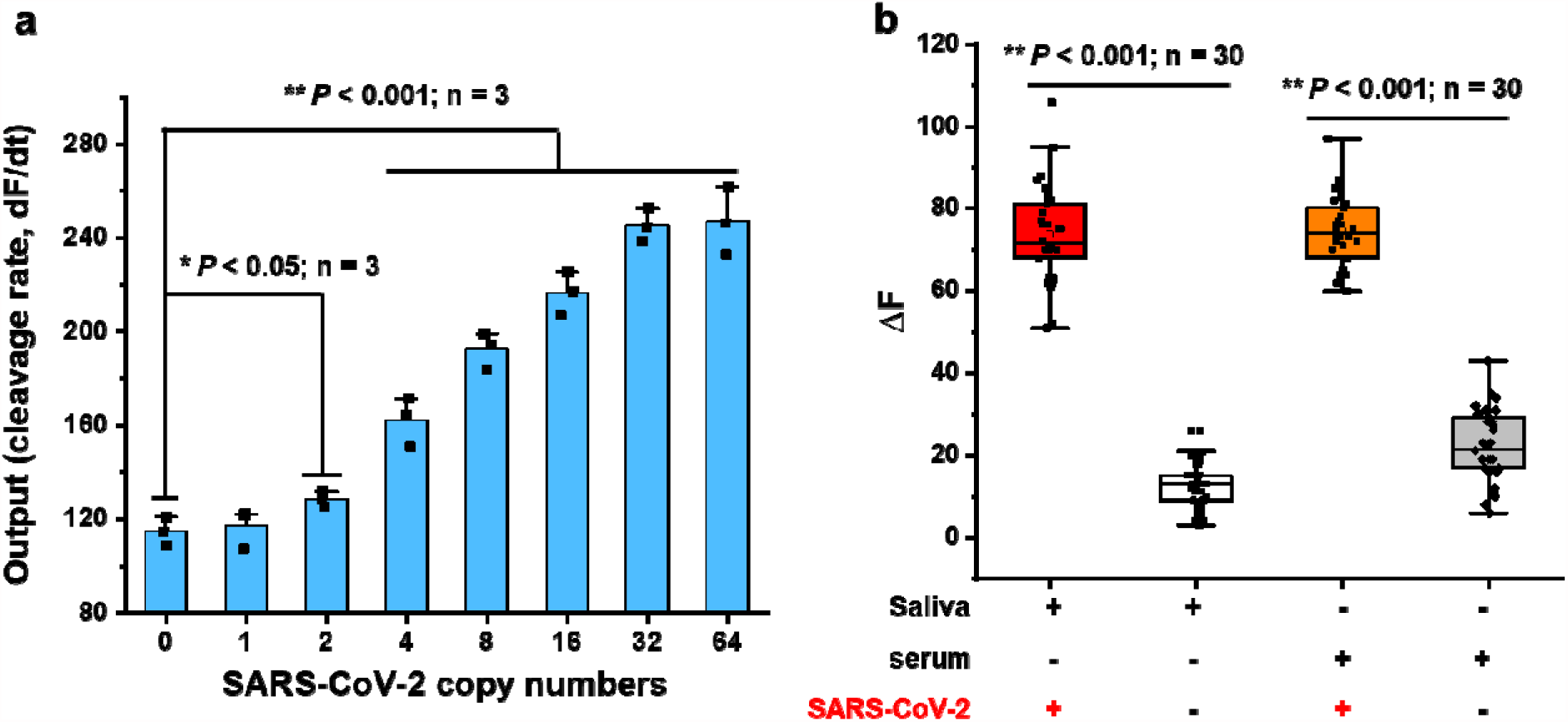
Detection of clinical inactivated SARS-CoV-2 virus. **(a)** Detection of clinical inactivated SARS-CoV-2 virus detected by CaT-Smelor-Covid.v3 platform. Data and SD are the means of three independent replicates. **(b)** Single blind test results of human saliva or serum samples spiked with or without inactivated SARS-CoV-2. ΔFI: fluorescence intensity increased in 1 min. The test results showed that the correct detection rate was 100%.

**Figure 5.**
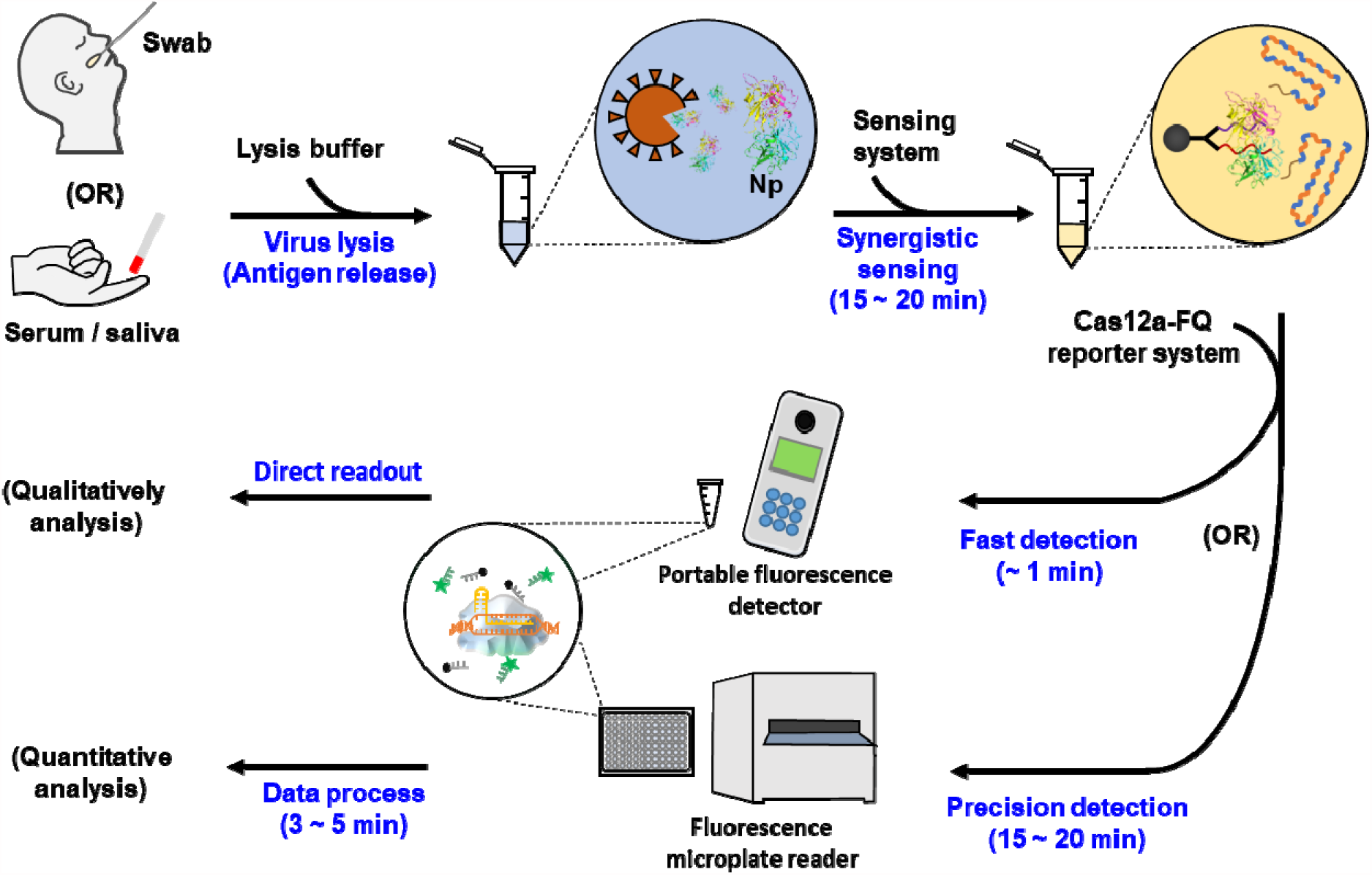
Workflow chart of sample detection. Swab samples and serum samples were collected and then treated to release the Np. Next, lysed solution was incubated with the biosensor system. Finally, the fluorescence intensity of released HyDNA was detected with a portable fluorometer or microplate reader.

### 2.5 Perspectives on the state-of-the-art, signal amplifiable CRISPR/Cas12a aptamer-mediated protein detection platform

Comparison our new Cas12a-aptamer-based protein detection platform with other previously reported SARS-CoV-2 virus detection methods indicated that CaT-Smelor-Covid.v3 was more sensitive and accurate in rapid antigen detection. The results of our method are comparable to those of the gold-standard RT-qPCR assays and other CRISPR-based nucleic acid detection approaches ^[8, 10]^ (Table 1). In addition, our rational optimization procedures applied to the CaT-Smelor platform successfully boosted the signal about 200-fold. Given its rational and modular configuration, our system after optimization should be widely applicable in many protein detection-based applications.

**Table 1.**
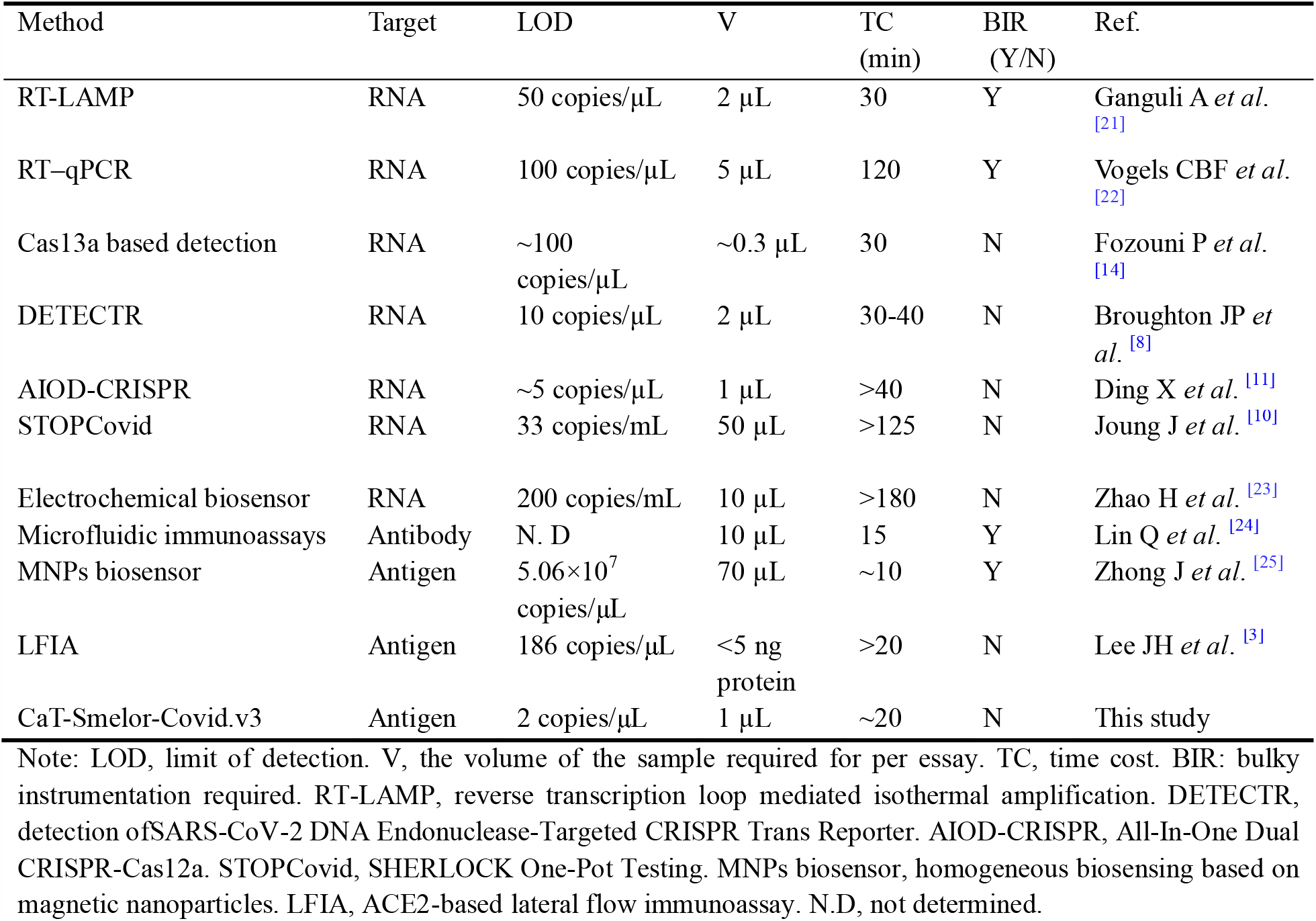
Comparison of reliable SARS-CoV-2 detecting methods.

## 3. Discussion

In parallel to the nucleic acid detection systems DETECTR, SHEROLOCK, and STOPCovid, the combination of CRISPR/Cas12a and aptamers represents a new detection paradigm for antigen detection ^[26]^. Coupling with aptamers dramatically broadens the detection scope of CRISPR/Cas12a ^[12, 17]^. However, how to stably amplify the signals sensed by aptamers and then translate the amplified signal into Cas12a cleavage activity has been an unresolved problem. Because of the lack of stable amplification of the signal sensed by the aptamer, this new detection paradigm has been unable to match the sensitivity of DETECTR and SHEROLOCK. Similarly, the sensitivity of Np antigen detection through the combination of CRISPR/Cas12a and aptamer in this study (LOD of 32 fM; ca. 193 copies/μL; Fig. 1, Supplementary Fig. 4) was much lower than the nucleic acid detection systems DETECTR, SHEROLOCK, and STOPCovid, although the performance of CaT-Smelor-Covid.v1 was comparable to those of the previously reported immunology-based methods (186 viral copies/μL) ^[24]^, Therefore, novel strategies were needed to dramatically enhance the sensitivity of this new detection paradigm. Furthermore, our results represent the general challenges of low sensing efficiency and detection sensitivity in the detection of target analytes through a combination of CRISPR/Cas12a and aptamers. Therefore, the main goal of this study was to address these problems and to provide a general solution for this detection paradigm.

In this study, multiple CRISPR/Cas12a trigger dsDNAs (up to 10 copies) were designed and constructed in tandem in a single DNA fragment (Fig. 2). The use of these trigger dsDNAs significantly amplified the output fluorophore signal. Of note, when the trigger-dsDNA copy number was increased from one (HyDNA) to 10 (HyDNA10), the detection sensitivity also linearly increased approximately 10-fold (from 32 fM to 3.48 fM). Warranting the stabilized output signal and maximization of the S/N, we designed both the crRNA and trigger dsDNA sequences on the basis of the phage genome-derived prediction site ^[27]^, which ensure optimal and robust performance of our CRISPR/Cas12a-based signal output system. In addition, the intersequence region between each trigger dsDNA was optimized. On the basis of our results, we believe that this well-characterized multiple trigger dsDNA tandem element could be used as a general tool in other CRISPR/Cas12a and aptamer-based detection systems, such as CaT-Smelor (16), in applications beyond Np detection.

For most aptamers, the affinity toward the corresponding target is usually at the nanomolar level (i.e., dissociation constant, *K*_*D*_), and further increasing the binding affinity by systematic evolution of ligands by exponential enrichment (SELEX) may be difficult ^[28]^. However, the *K*_*D*_ of the aptamer directly determines the sensing efficiency and consequently affects the detection sensitivity. Therefore, innovating the aptamer-based sensing module for a more sensitive response to antigen, independently of the affinity between aptamer and antigen, should be of general interest for all aptamer-based detection systems. In this study, we used a Y-shaped DNA and two aptamers, A48 and A61, recognizing different epitopes of the antigen, which were designed to sense the Np (Fig. 3). A significant synergistic sensing effect was observed, in which the binding of A48 to Np facilitated the binding of A61 and Np, and vice versa. According the response curve, the Hill coefficient reached 1.52, thus indicating a positive synergistic effect between A48 and A61 ^[29]^. By combining the multiple trigger dsDNA and synergistic sensing strategies, our system achieved an LOD for Np detection of 0.17 fM (Supplementary Fig. 10). Therefore, our dual aptamer synergistic sensing strategy, independently of binding affinity, not only increased the affinity limit of aptamers but also further improved the detection sensitivity (from 3.48 fM to 0.17 fM). Moreover, the synergistic effects could be applied to the detection of any other antigen with two aptamers that recognize different epitopes of the antigen.

The output of the ssDNA cleavage activity of Cas12a can detected through many convenient methods, e.g., lateral-flow ^[30]^. Here, we used a commercially available portable fluorescence detector to demonstrate the merits and potential of our method in clinical diagnosis. Saliva and serum spiked with different concentrations of inactivated SARS-CoV-2 were used to mimic clinical samples, and the positive detection rate reached 100% (Fig. 4b). Despite the limitations of the experimental conditions, and the absence of clinical trials, we believe that our results confirmed the practicability of our detection approach. In addition, compared with the existing antigen detection methods, our method had significant advantages in that the sensitivity was greatly improved, and the testing is fast in time-spent and low in cost (Table 1). The abovementioned advantages of CRISPR/Cas-based detection systems also extended to any antigen detection with our strategy.

## 4. Conclusion

Consequently, by focusing on the issues of signal amplification and sensing efficiency in the CRISPR/Cas12a and aptamer-based detection paradigm, we not only developed a PCR-free multiple trigger dsDNA tandem-based signal amplification strategy for high performance CRISPR/Cas-based detection but also designed a dual aptamer synergistic sensing strategy for highly efficient sensing. Integrating these two aspects, we achieved the first reported ultrasensitive, fast, stable, high-performance antigen detection using a CRISPR/Cas12a and aptamer-based detection paradigm. Given that aptamers recognizing different fragments of variable proteins (for example, multiple epitopes of antigens) can be generated by SELEX, we believe that the innovative strategy used in this study may mark the beginning of a wide range of applications in the development of detection methods for antigens and other target molecules.

## 5. Experimental Section

### Materials and reagents

Aptamers and primers were chemically synthesized by Beijing Tsingke Biotech Co. Ltd (Beijing, China) (Table S1). Beaver Beads™ streptavidin (1 μm average diameter) was purchased from Beaver (Suzhou, China). SARS-CoV-2 Np was purchased from Sino Biological Inc (Beijing, China). Human serum and other chemicals for buffers and solvents were purchased from Sigma-Aldrich, Inc. (Shanghai, China). UltraPure DNase/RNase-free Distilled Water, HiScribe T7 Quick High Yield RNA Synthesis Kit, *Taq* 2× Master Mix, standard *Taq* buffer and RNase inhibitor were purchased from New England BioLabs Inc. (Ipswich, UK). The TIANquick Maxi Purification Kit was purchased from Tiangen Biotech Co. Ltd (Beijing, China). The RNA Clean & Concentrator™-5 was purchased from Zymo Research (Irvine, CA). All chemicals and reagents used were of analytical grade and were prepared with deionized water (18 MΩ cm) from a Milli-Q^®^ Water Purification System (Millipore, Billerica, MA, USA). The portable fluorometer was purchased from Thermo Fisher Scientific Inc.

### Cas12a protein expression and purification

Cas12a (LbCas12a) from Lachnospiraceae bacterium ND2006 was expressed and purified as described in our previous study. Briefly, *Escherichia coli* BL21(DE3) was transformed with the pET28TEV-LbCas12a expression plasmid and grown overnight in Luria-Bertani medium at 37°C with shaking at 150 rpm until the exponential growth phase. Protein expression was then induced with 100 µM isopropyl-β-D-thiogalactoside, and the cells were cultured at 16°C for 12 h. The cells were harvested by centrifugation at 5,000 × *g*. For protein purification, the cell pellets were resuspended in lysis buffer (50□mM Tris-HCl [pH 7.4], 200□mM NaCl, 2□mM dithiothreitol, and 5% glycerol) supplemented with EDTA-free protease inhibitors (Sigma-Aldrich, Roche Diagnostics), and then lysed with ultrasonication. The lysate was loaded onto a HisTrap FF column (GE Healthcare) and washed with an imidazole concentration gradient. The peak fractions were collected and desalted with dialysis. The solution was then loaded onto a HiTrap Q HP column (GE Healthcare), and the peak fractions were collected, pooled, and concentrated. The concentrated solution was loaded onto a HiLoad 16/600 Superdex 200□pg column for fast protein liquid chromatography (AKTA Explorer 100; GE Healthcare). The gel filtration fractions were analyzed with SDS-PAGE, and the protein concentrations were determined with the Bradford method. The purified product was dissolved in storage buffer (20□mM Tris-HCl [pH 7.5], 1□M NaCl, and 50% glycerol) and stored at -80°C until use.

### HyDNA, crRNA, and Y-DNA preparation

The oligonucleotides used in this work are listed in Table S1. To prepare the template for HyDNA amplification, a DNA sequence with ten copies of Cas12a trigger dsDNA was synthesized. Then, C3 modified forward primers (com1F, com2F, com3F, com4F, com5F, com6F, com7F, com8F, com9F, com10F, com11F, com12F, com13F, com14F, com15F, com16F, com17F, com18F, com19F, and com20F) combined with reverse primers (dsDNA-R1, dsDNA-R2, dsDNA-R3, dsDNA-R4, dsDNA-R5, dsDNA-R6, dsDNA-R7, dsDNA-R8, dsDNA-R9, and dsDNA-R10) were amplified by using the synthesized template DNA to produce the HyDNAs containing Cas12a trigger dsDNA and different com-ssDNA sequences (ssDNA complementary to part of the sequence of the aptamer) ^[31]^. The PCR products were generated with *Taq* 2× Master Mix. The PCR conditions were 1 min at 95°C for activation followed by 30 cycles at 95°C for 30 s for denaturation, 55°C for 30 s for annealing, 68°C for 30 s for elongation, and a final cycle at 68°C for 10 min for final elongation. The HyDNAs were purified with a TIANquick Maxi Purification Kit.

To prepare the template for crRNA (crRNA1, crRNA2, crRNA3, crRNA4, crRNA5, crRNA6, crRNA7, crRNA8, crRNA9, and crRNA10) synthesis, paired oligonucleotides containing a T7 priming site (Table S1) were synthesized and annealed in 1× *Taq* DNA Polymerase PCR Buffer. The crRNAs were then transcribed with a HiScribe™ T7 Quick High Yield RNA Synthesis Kit and purified with RNA Clean & Concentrator™-5.

To obtain the Y-DNA, three oligonucleotides (Y1, Y2, and Y3) were designed. Each oligonucleotide sequence was complementary to the other two oligonucleotide sequencess. For the preparation of the Y-DNA based biosensor, the produced Y-DNA contained 25 bases as a sticky end on the two arms (Y1 and Y2) and a biotin labeled poly A end on the Y3 arm. For construction of Y-DNA, the same molar amounts of Y1, Y2, and Y3 were mixed with *Taq* DNA Polymerase PCR buffer. Then the mixture was incubated at 95°C for 2 min, quickly cooled to 65°C, incubated at 65°C for 2 min, cooled to 60 °C at a rate of -1°C per min, incubated at 60°C for 2 min, cooled to 20°C at a rate of -0.1°C per 30 s, and finally cooled to 16°C ^[32].^ The resulting HyDNAs and crRNAs were quantified with a NanoDrop 2000 spectrophotometer (Thermo Fisher Scientific). The HyDNAs and Y-DNA were stored at 4°C. The crRNAs were stored at -80°C. Throughout all experiments, RNase-free materials and conditions were used.

### Biosensor configuration and sensing

To construct the one aptamer-based biosensor, MB, biotin labeled aptamer, and HyDNA were used. First, 20 μL of 10 mg/mL MB was washed with 1× PBS containing Tween 20 (PBST) buffer to remove the residual NaN_3_ protection solution. Then 30 μL of 10 μM biotin labeled aptamer in 1× PBST buffer was added to the MB solution and mixed well for 30 min at room temperature. The aptamer-coated MB were then washed twice with buffer to remove excess biotin labeled aptamer. Then the HyDNA (0.5 μM, 20 μL) was added to the solution and mixed well at room temperature for 20 min. After the sample was washed three times with buffers to remove excess HyDNAs, the MB-aptamer-HyDNA complex was dispersed in 1 mL of 1× PBST buffer. The MB in each 20 μL aliquot of this solution were used in this work.

To construct the dual aptamer-based biosensor, MB, biotin labeled Y-DNA, adapter1-A48, adapter2-A61 ^[19]^, com10-HyDNA and com11-HyDNA were used. First, 20 μL of 10 mg/mL MB was washed with 1× PBST buffer to remove the residual NaN_3_ protection solution. Then 300 μL of 1 μM biotin labeled Y-DNA was added to the MB solution and mixed well for 30 min at room temperature. The MB were then washed twice with buffer to remove excess Y-DNA. The adapter1-A48 (10 μM, 30 μL) and adapter2-A61 (10 μM, 30 μL) were added to the solution and mixed well at room temperature for 20 min. The MB were washed twice with buffer to remove excess aptamers. The HyDNAs (0.5 μM, 10 μL) were added to the solution and mixed well at room temperature for 20 min. After the sample was washed three times with buffers to remove excess HyDNAs, the MB-YDNA-aptamer-HyDNA complex was dispersed in 1 mL of 1× PBST buffer. The MB contained in each 20 μL aliquot of this solution were used in this work.

To assess the feasibility of the platform, we incubated samples with different concentrations of Np with the biosensor at room temperature for 20 min. Each solution was then separated with a magnetic rack, and 10 µL of supernatant was transferred into a new centrifuge tube for quantification with the Cas12a-based FQ-labeled reporter system.

### Quantification of HyDNA with the CRISPR-Cas12a reporting system

In the CRISPR-Cas12a cutting system, NEB CutSmart^®^ Buffer was used as the reaction buffer. For the collateral cleavage assay, an equimolar ratio of Cas12a and crRNA was premixed with the FQ-labeled reporter in NEB CutSmart^®^ Buffer and then distributed in a 384-well plate for the subsequent experiment. The diluted supernatant containing the HyDNA was added to the reporter system, and the fluorescence signal was detected every 1.5 min with an EnSpire™ Multimode Plate Reader (PerkinElmer, Inc., USA) at an excitation wavelength of 480 nm and an emission wavelength of 520 nm.

### Analysis of samples

An inactivated SARS-CoV-2 sample was obtained from Dr. Bi and was quantified previously by using gene N primers. To demonstrate the analytical reliability and practical application of the biosensing platform, we tested swab samples spiked with different concentrations of inactivated SARS-CoV-2. First, 10 µL inactivated SARS-CoV-2 sample was incubated in 10 µL lysis buffer for 10 minutes and serially diluted in half to obtain different concentrations of lysed samples. Each diluted sample (2 µL) was added to 18 µL of the biosensor system and incubated for 20 min. To construct the calibration curve, we detected the HyDNA in the supernatant with the Cas12a-based FQ-labeled reporter system. The changes in the fluorescence intensity over time were measured, and the slope of the fluorescence curve in the linear region between 0 and 8 min (normally) represented the ssDNA trans-cleavage rate of CRISPR–Cas12a. The linear relationship between the trans-cleavage rate (or slope) and the SARS-CoV-2 standard concentration was obtained.

To demonstrate that SARS-CoV-2 screening with our platform would be possible outside of laboratory settings, we used a portable fluorometer (Qubit™ 4 Fluorometer) to quantify the fluorescence signal generated by the detection assay. The unactivated SARS-CoV-2 sample was diluted to three to five copies/μL, and Np was released with lysis buffer. The extracted Np sample (2 µL) was added to 20 µL of the biosensor system and incubated for 20 min. Then the HyDNA in the supernatant was detected with the Cas12a-based FQ-labeled reporter system. The fluorescence signal increased in 1 min was detected with the portable fluorometer at an excitation wavelength of 470 nm and an emission wavelength of 510–580 nm.

### Data analysis

All experiments and assays were repeated at least three times. The data in all figures are expressed as means ± SE. Microsoft Excel 2016, Origin 8, and GraphPad Prism version 6.0 were used for data analysis.

## Data Availability

All data referred to in the manuscript is available from the corresponding authors.

## Supplementary data

Supporting Information is available from the Wiley Online Library or from the author.

## Acknowledgements

This work was supported by the National Key R&D program of China (2020YFA0907800), the National Natural Science Foundation of China (31922002, 31720103901, 31772242 and 31870040), the 111 Project (B18022), the Fundamental Research Funds for the Central Universities [22221818014], the Youth Innovation Promotion Association CAS (Y202027) to W.W and the Open Project Funding of the State Key Laboratory of Bioreactor Engineering.

## Conflict of Interest

The authors declare no conflict of interest.

## Author contributions

Lixin Zhang, Gao-Yi Tan and Weishan Wang conceived and supervised the project. Xiangxiang Zhao, Zhengduo Wang and Bowen Yang designed and performed the experiments. Xiangxiang Zhao and Gao-Yi Tan analyzed the data. Zhengheng Li, Xuekui Xia and Xiangyin Chen participated in this work. Yuhai Bi provided inactivated virus samples. Gao-Yi Tan, Weishan Wang, Yaojun Tong and Lixin Zhang wrote the manuscript.

## Supplementary information

## Supplementary Figures

**Supplementary Figure 1.**
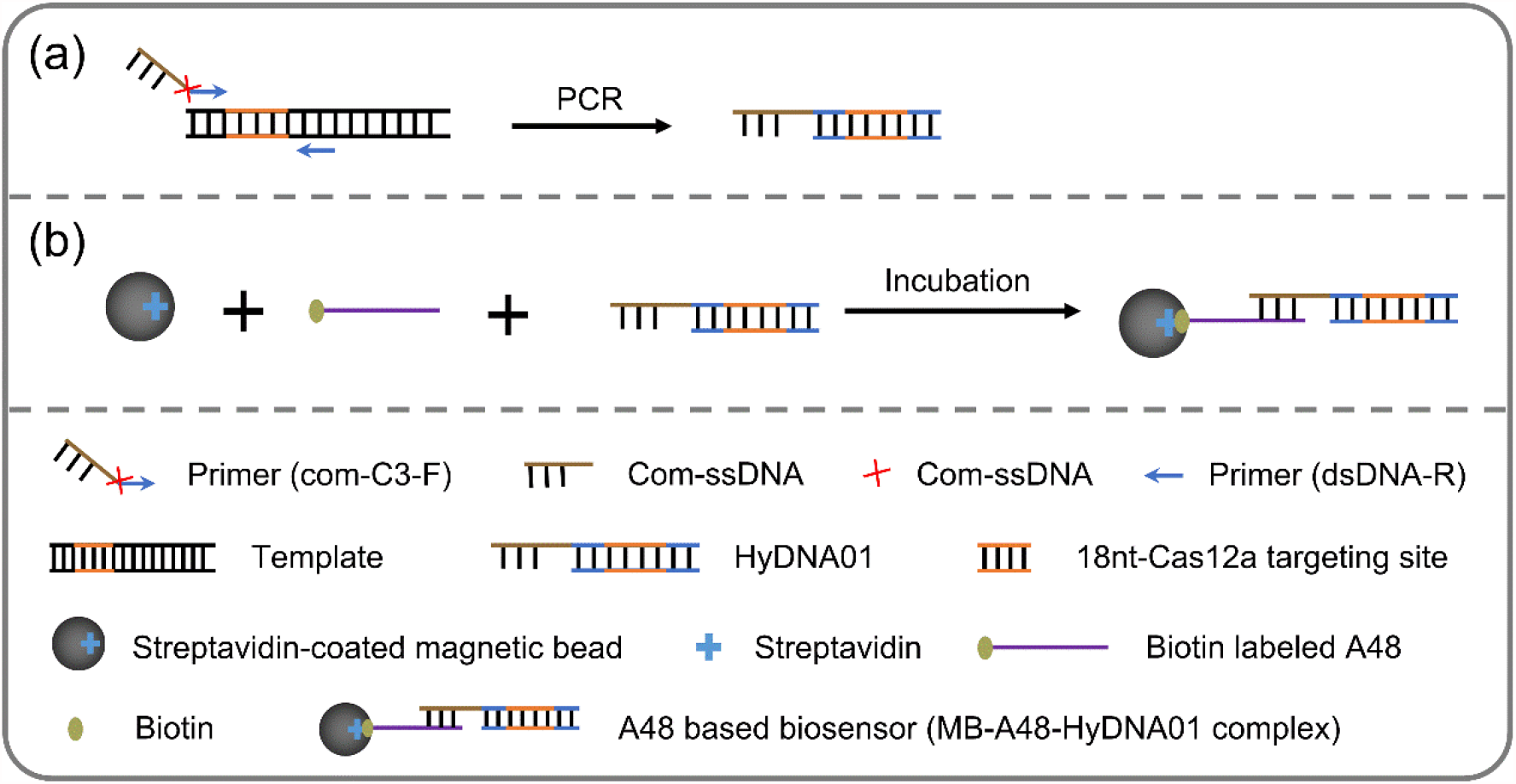
Illustration of the assembly of the CaT-Smelor-Covid.v1 platform. **(a)** Production of HyDNA01 by PCR using primer that contains C3 spacer. **(b)** Assembly of A48 based biosensor by mixing MBs, biotin labeled A48 and HyDNA01.

**Supplementary Figure 2.**
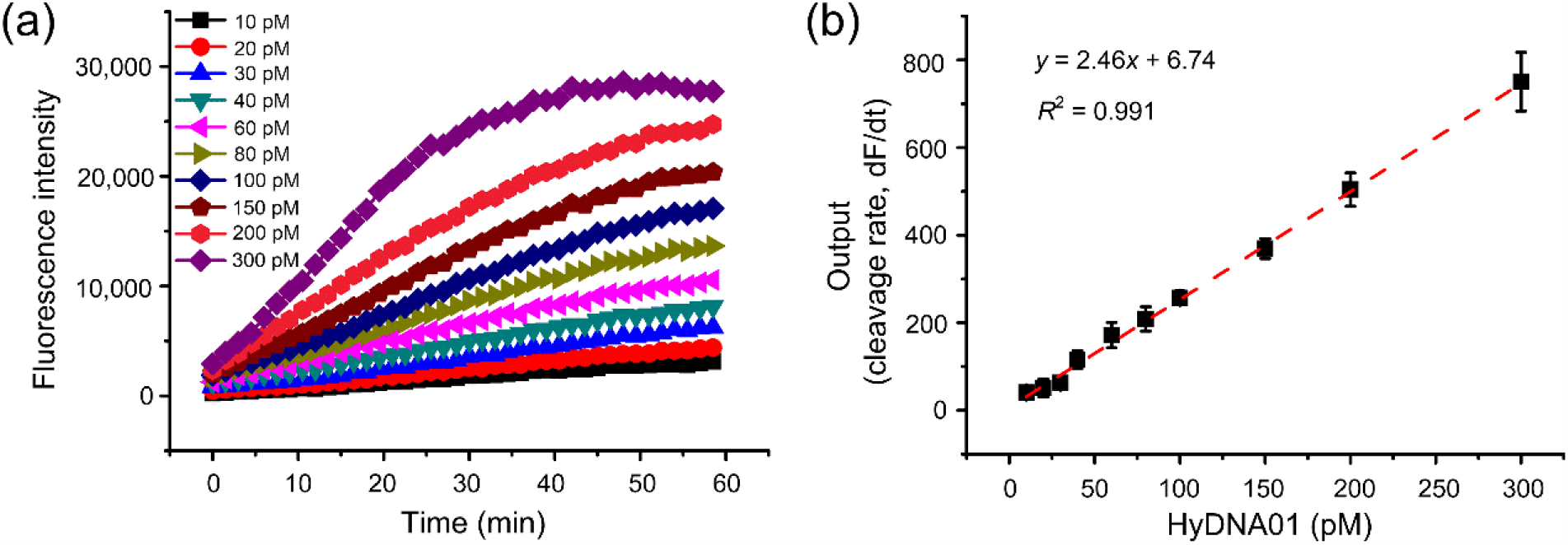
Effects of different concentration of activator HyDNA01 on the ssDNA trans cleavage activity of CRISPR/Cas12a. **(a)** Plot of the fluorescence level against the concentration of HyDNA01. **(b)** The linear relationship between the activator HyDNA01 concentration and the slope of the fluorescence intensity (cleavage rate, dFI/dt) of CRISPR/Cas12a. The changes in the fluorescence intensity with time were measured, and the slope of the fluorescence curve in the linear region between 0 and 8 min (normally) represented the ssDNA trans-cleavage rate of CRISPR/Cas12a. Linearity range of the calibration curve was 10–300 pM (*R*^2^ = 0.991). Data were the means and SE of three independent replicates.

**Supplementary Figure 3.**
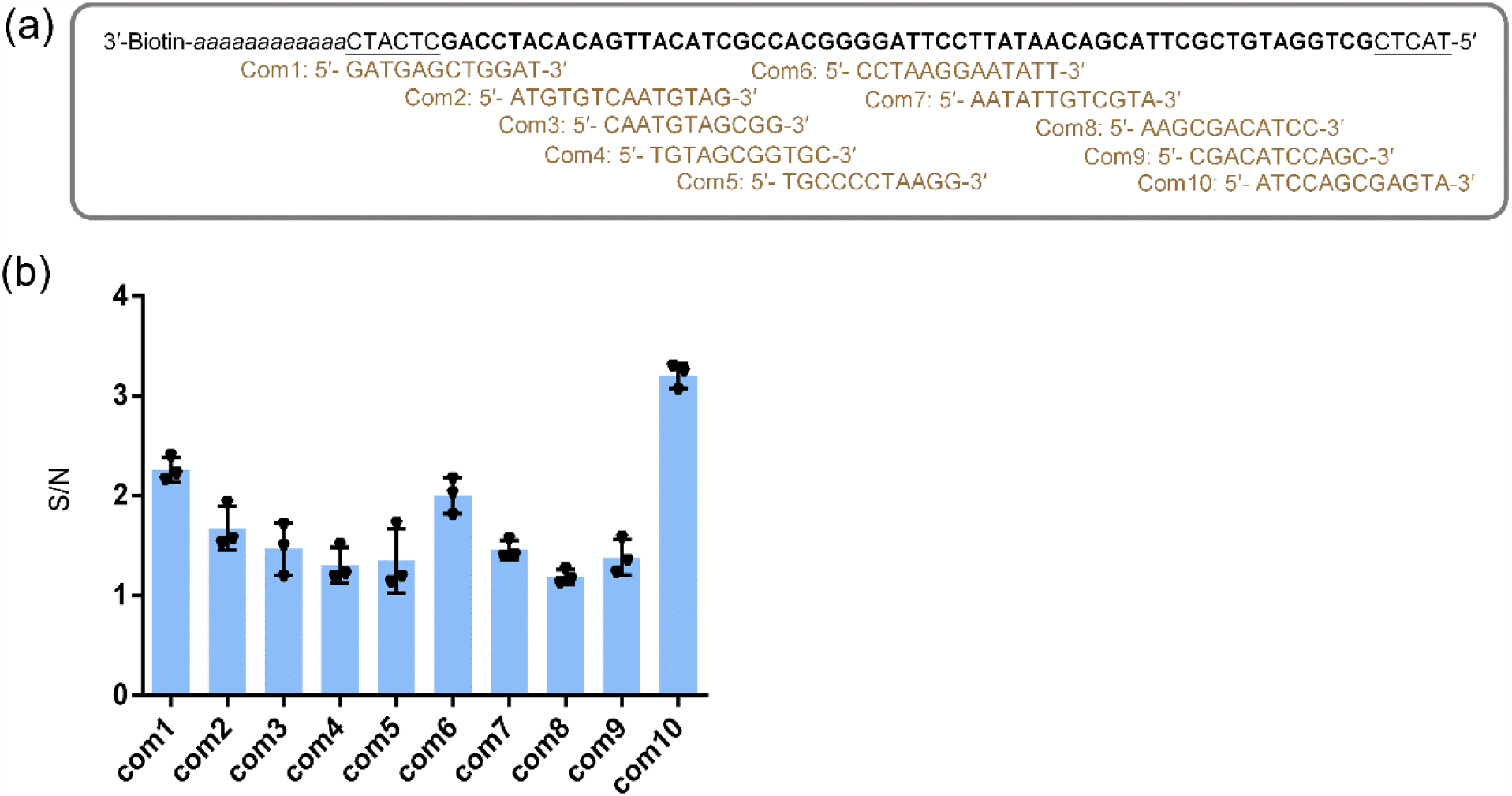
Signal-to-noise ratio optimization of aptamer A48 by com-site selection. **(a)** Schematic diagram of Com-site candidates. The sequence marked bold indicates the sequence of A48. The underline sequences indicate the additional sequences added at both ends of the aptamer. The poly A indicates linker sequence. Com1, Com2, Com3, Com4, Com5, Com6, Com7, Com8, Com9 and Com10 are designed Com-site candidates, which are complement with different regions of biotin labeled aptamer sequence, respectively. **(b)** Signal-to-noise ratio of com-site candidates. The released HyDNA01 was detected. Since com10 gave the highest S/N ratio, it was chosen to configure the HyDNA biosensor. Data were the means and SE of three independent replicates.

**Supplementary Figure 4.**
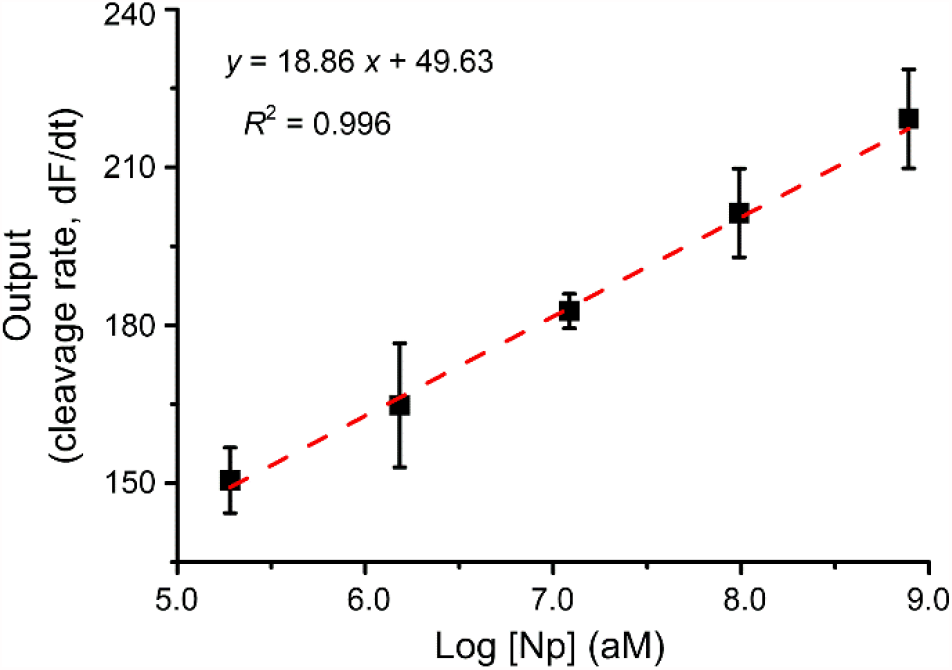
Gathering the output data of the CaT-Smelor-Covid.v1 platform. The linear relationship between the Np concentration and the slope of the fluorescence intensity (cleavage rate, dFI/dt) of CRISPR/Cas12a. Data were the means and SE of three independent replicates.

**Supplementary Figure 5.**
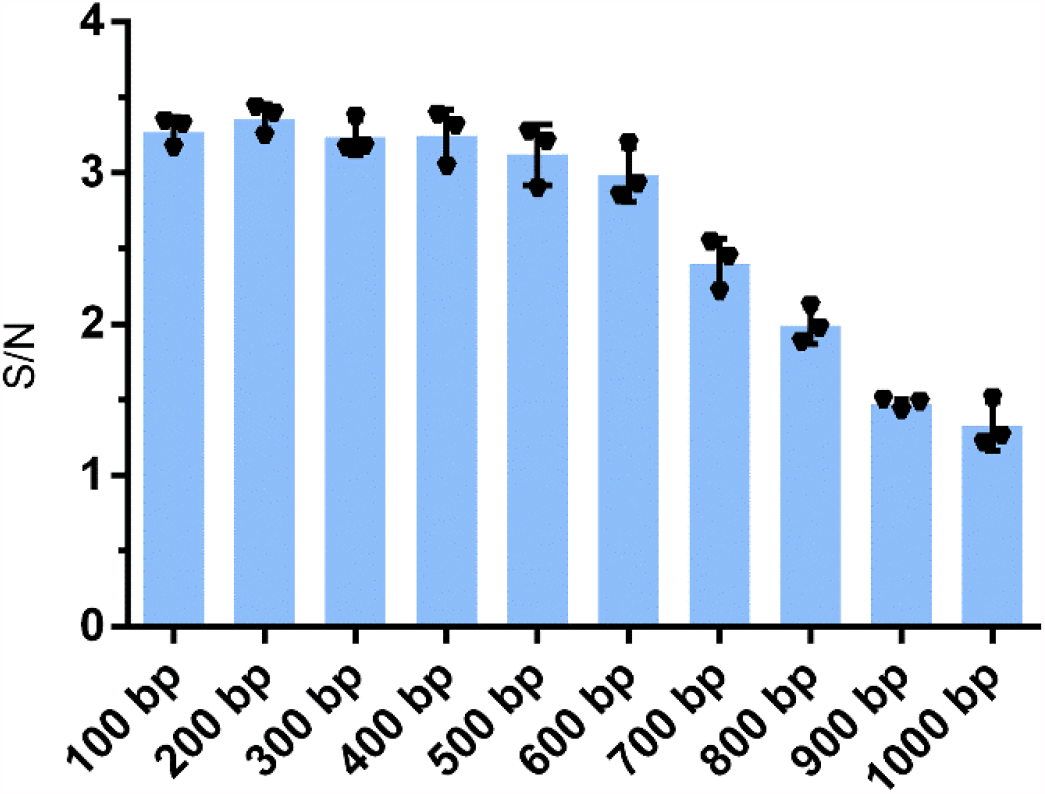
Signal-to-noise ratio of different lengths of double-stranded DNA in HyDNA. The signal-to-noise ratio of sequences longer than 600bp was obviously decrease. Data were the means and SE of three independent replicates.

**Supplementary Figure 6.**
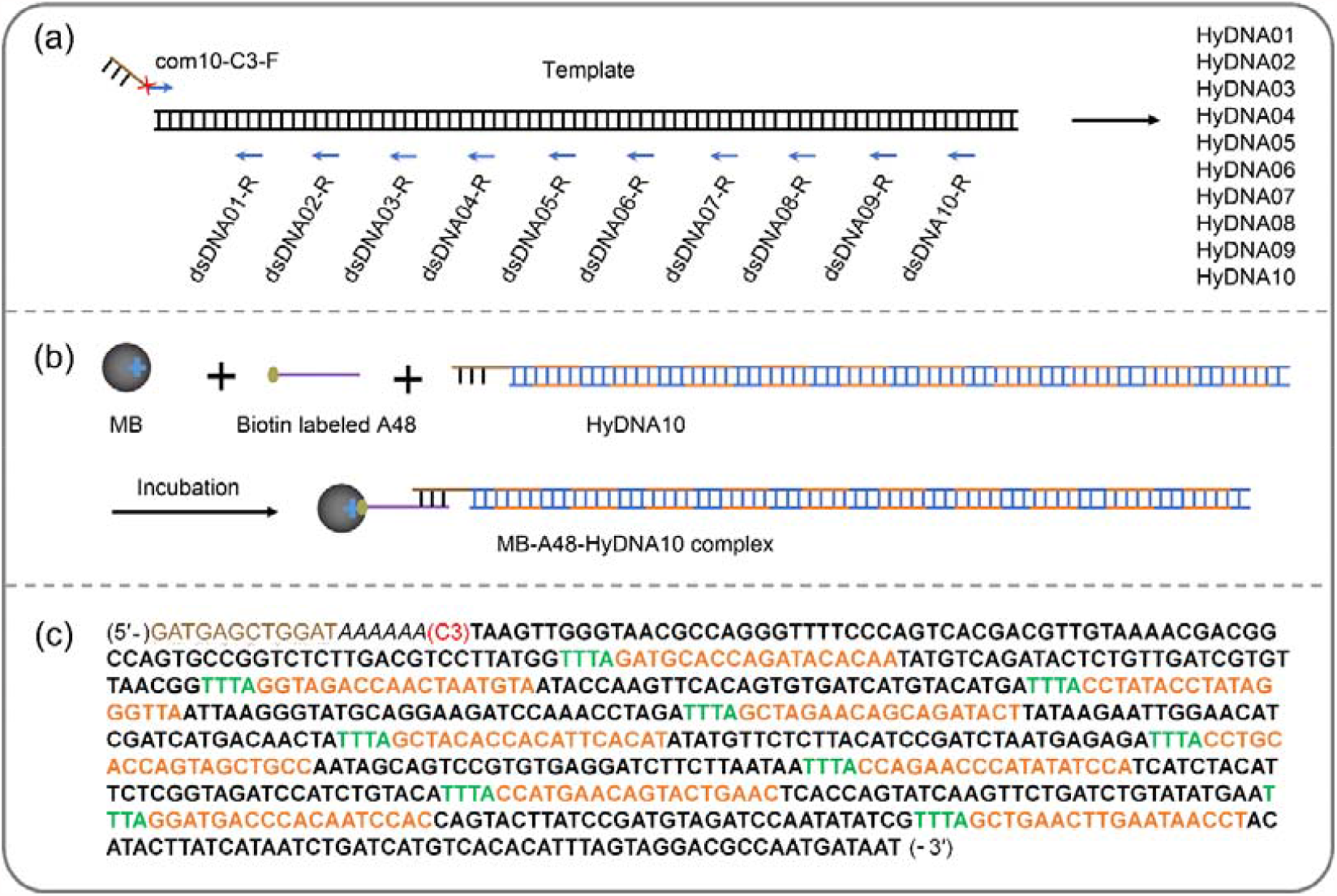
Illustration of the assembly of the CaT-Smelor-Covid.v2 platform. **(a)** Preparation of HyDNAs containing different copy numbers (1–10) of Cas12a trigger dsDNAs. **(b)** Construction of MB-A48-HyDNA10. **(c)** Sequence of com10-HyDNA10. The Cas12a trigger dsDNA sequence was shown in bold. The PAM sites were marked as green. The Cas12a targeting sites were marked as orange. The sequence of com10 was marked as brown.

**Supplementary Figure 7.**
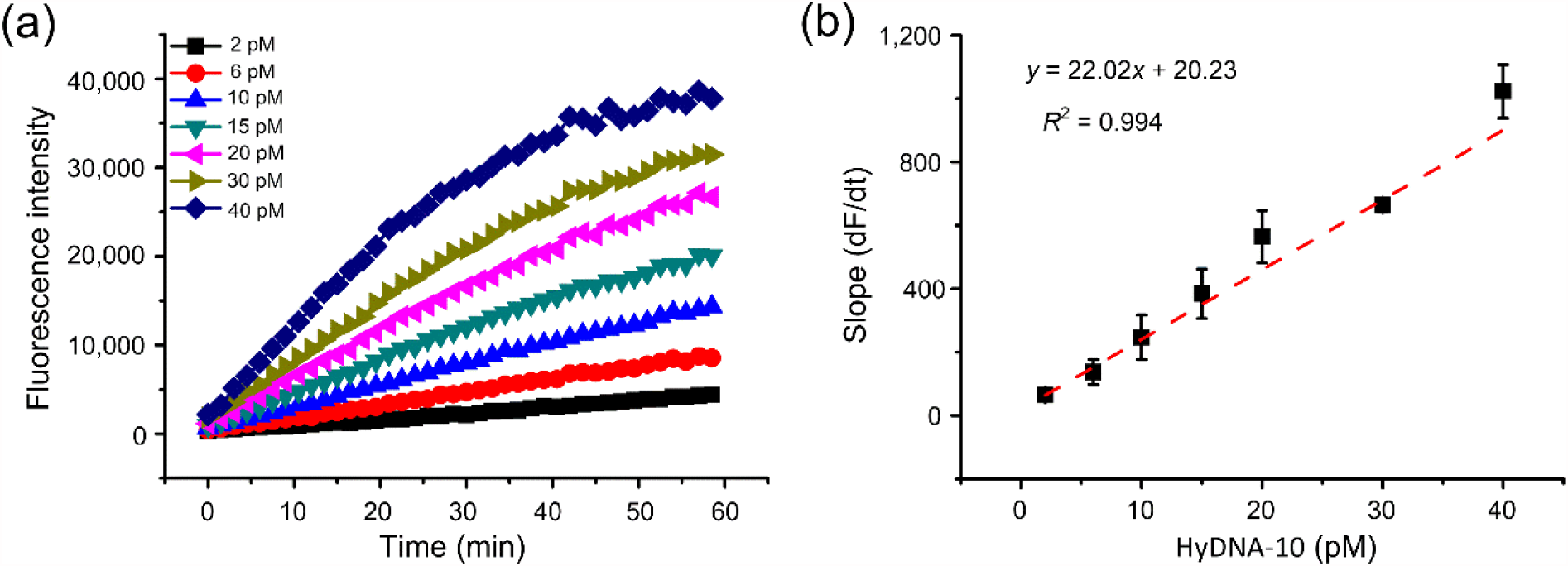
Effects of different concentration of activator HyDNA10 on the ssDNA trans cleavage activity of CRISPR/Cas12a. **(a)** Plot of the fluorescence level against the concentration of HyDNA10. **(b)** The linear relationship between the activator HyDNA10 concentration and the slope of the fluorescence intensity (cleavage rate, dFI/dt) of CRISPR/Cas12a. The changes in the fluorescence intensity with time were measured, and the slope of the fluorescence curve in the linear region between 0 and 8 min (normally) represented the ssDNA trans-cleavage rate of CRISPR/Cas12a. Linearity range of the calibration curve was 2–40 pM (*R*^2^ = 0.994). Data were the means and SE of three independent replicates.

**Supplementary Figure 8.**
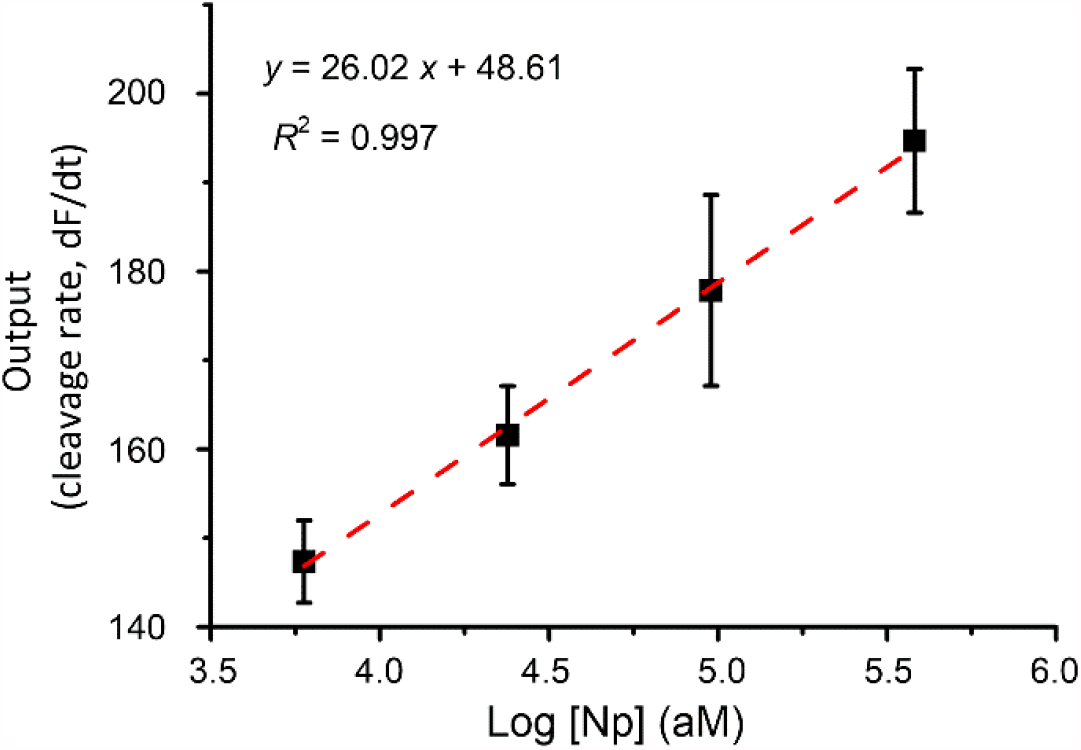
Gathering the output data of the CaT-Smelor-Covid.v2 platform. The linear relationship between the Np concentration and the slope of the fluorescence intensity (cleavage rate, dFI/dt) of CRISPR/Cas12a. Data were the means and SE of three independent replicates.

**Supplementary Figure 9.**
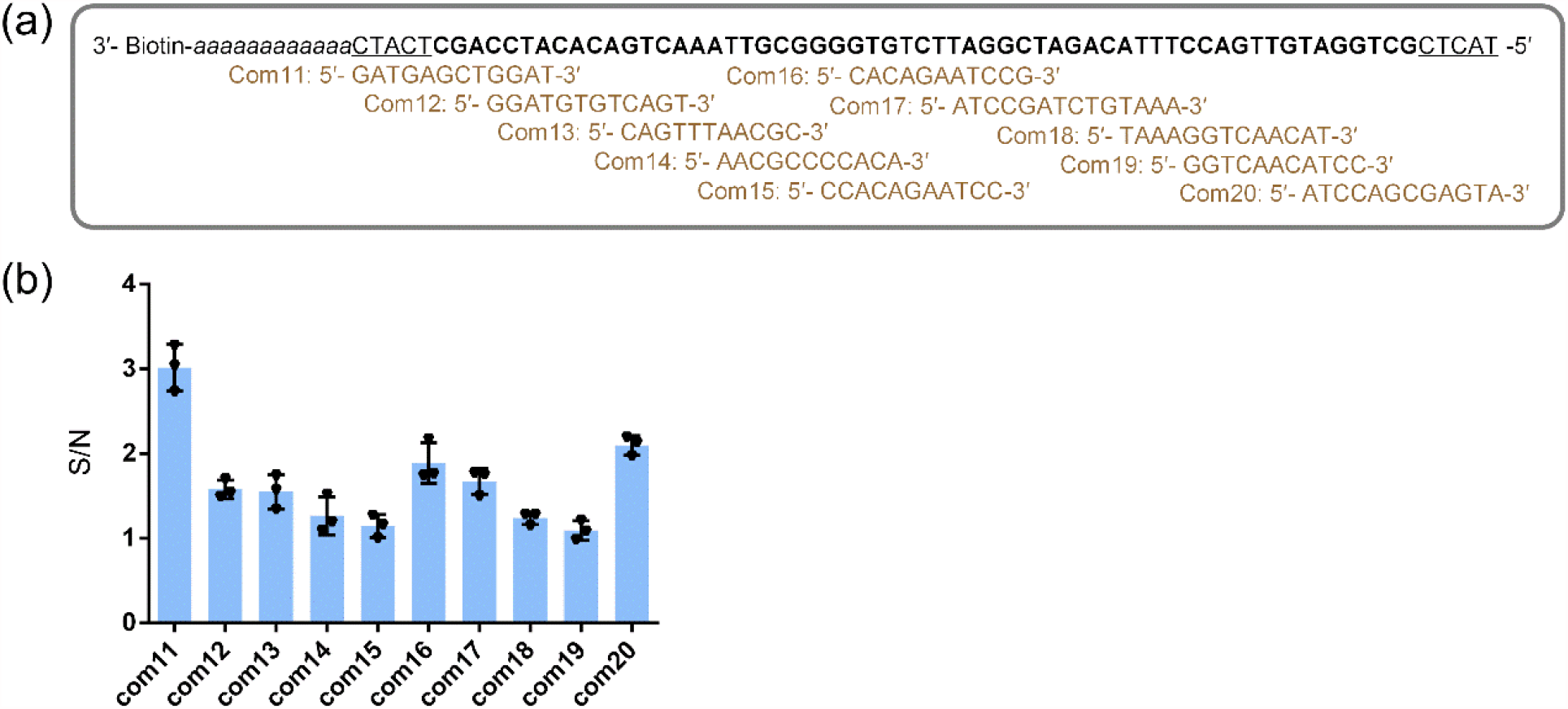
Signal-to-noise ratio optimization of aptamer A61 by com-site selection. **(a)** Schematic diagram of Com-site candidates. The sequence marked bold indicates the aptamer A61 sequence. The underline sequences indicate the additional sequences added at both ends of the aptamer. The poly A indicates linker sequence. Com11, Com12, Com13, Com14, Com15, Com16, Com17, Com18, Com19 and Com20 are designed Com-site candidates, which are complement with different regions of biotin labeled aptamer, respectively. **(b)** Signal-to-noise ratio of com-site candidates. Since com11 gave the highest S/N ratio, it was chosen to configure the HyDNA. Data were the means and SE of three independent replicates.

**Supplementary Figure 10.**
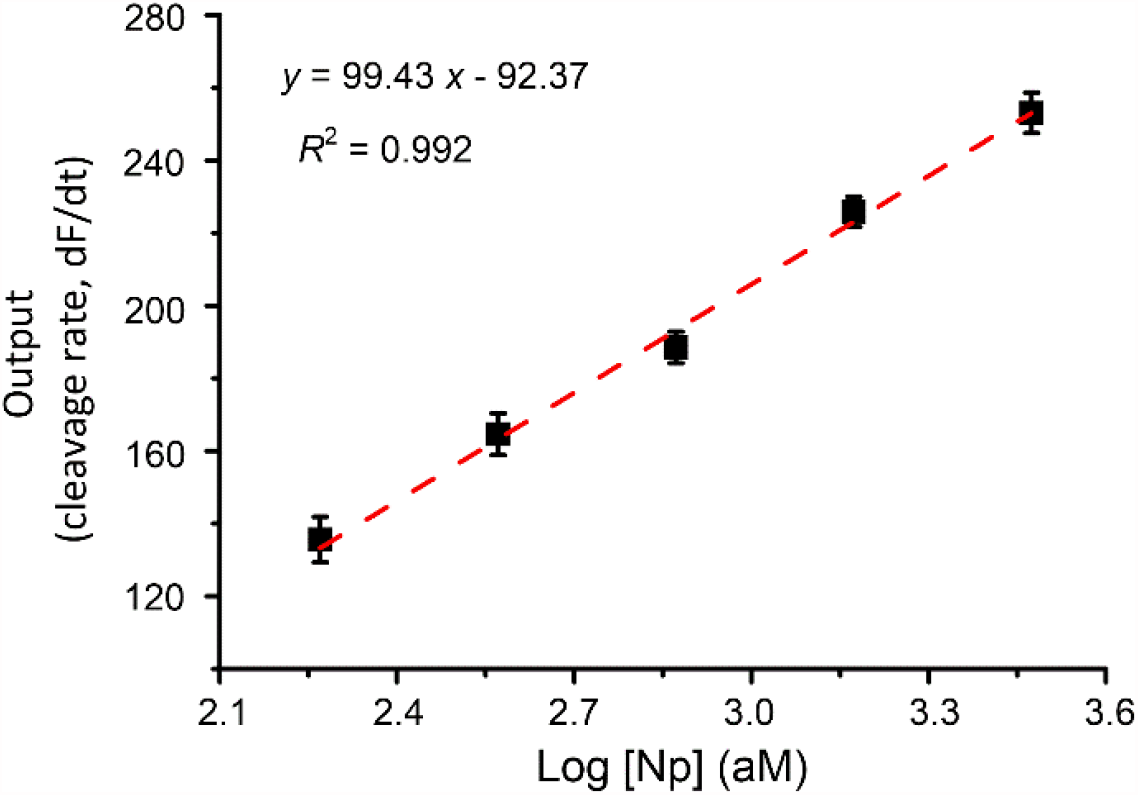
Gathering the output data of the CaT-Smelor-Covid.v3 platform. The linear relationship between the Np concentration and the slope of the fluorescence intensity (cleavage rate, dFI/dt) of CRISPR/Cas12a. Data were the means and SE of three independent replicates.

**Supplementary Figure 11.**
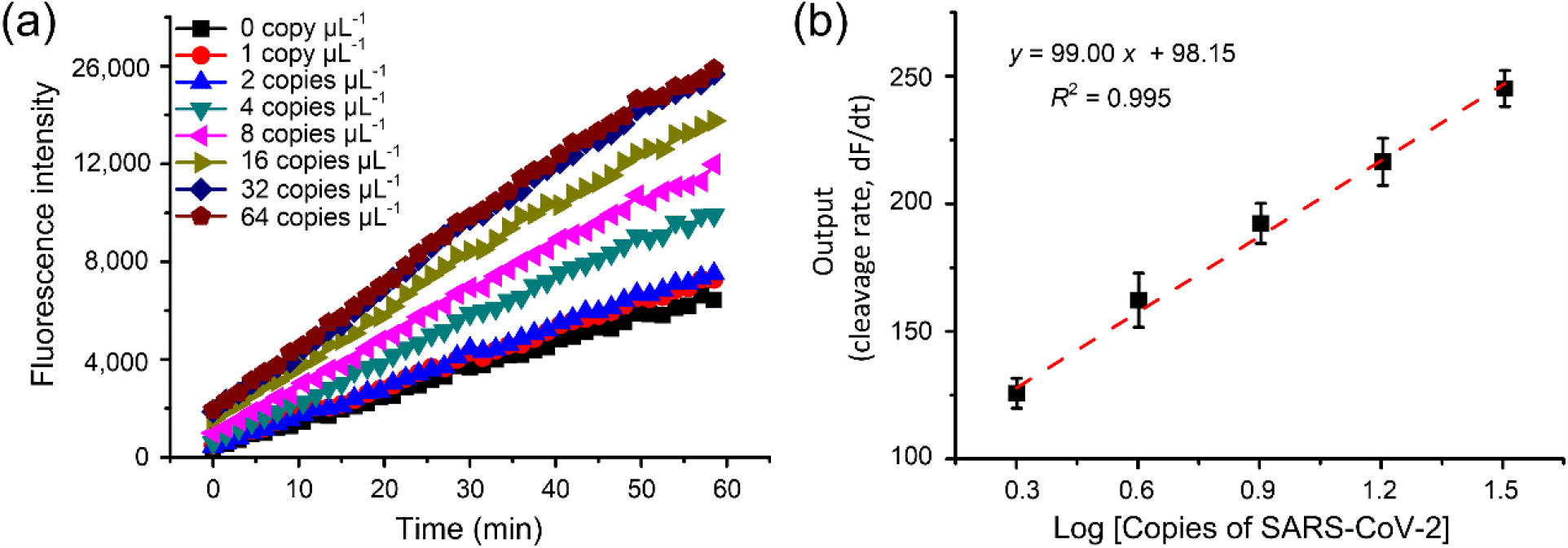
Standard curve line of inactivated SARS-CoV-2 samples detected by microplate reader in 384-well plates. **(a)** Plot of the fluorescence level against the concentration of SARS-CoV-2. **(b)** The linear relationship between the SARS-CoV-2 concentration and the slope of the fluorescence intensity (cleavage rate, dFI/dt) of CRISPR/Cas12a. The changes in the fluorescence intensity with time were measured, and the slope of the fluorescence curve in the linear region between 0 and 8 min (normally) represented the ssDNA trans-cleavage rate of CRISPR–Cas12a. Data were the means and SE of three independent replicates.

**Supplementary Figure 12.**
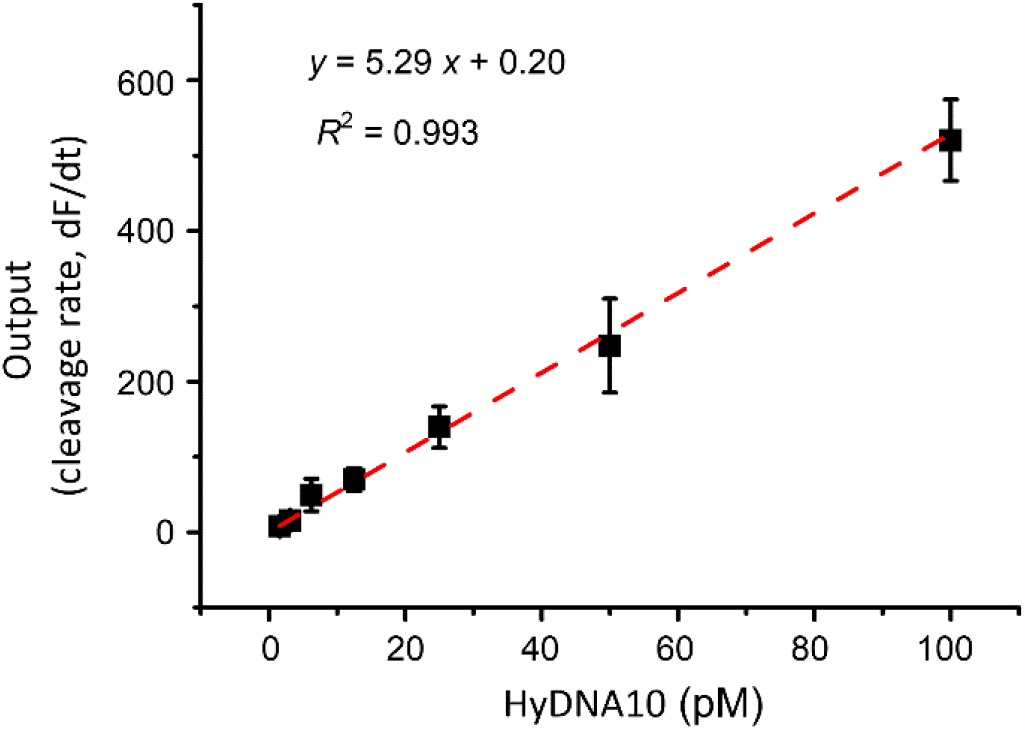
Effects of different concentration of activator HyDNA10 on Cas12a ssDNA trans cleavage activity detected by portable fluorometer. The linear relationship between the activator HyDNA10 concentration and the slope of the fluorescence intensity (cleavage rate, dFI/dt) of CRISPR/Cas12a. The changes in the fluorescence intensity with time were measured, and the slope of the fluorescence curve in the linear region between 0 and 60 s (normally) represented the ssDNA trans-cleavage rate of CRISPR/Cas12a. Data were the means and SE of three independent replicates.

**Supplementary Figure 13.**
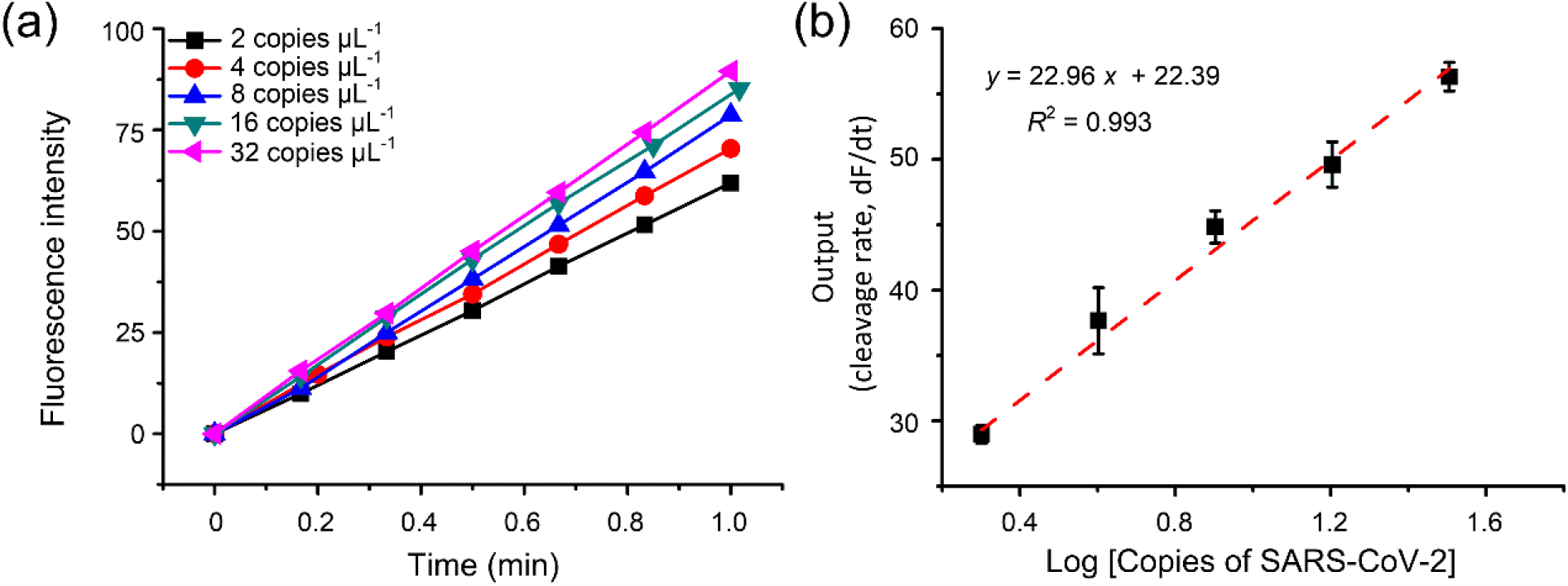
Standard curve line of inactivated SARS-CoV-2 samples detected by portable fluorometer. **(a)** Plot of the fluorescence level against the concentration of SARS-CoV-2. (b) The linear relationship between the SARS-CoV-2 concentration and the slope of the fluorescence intensity (cleavage rate, dFI/dt) of CRISPR/Cas12a. The changes in the fluorescence intensity with time were measured, and the slope of the fluorescence curve in the linear region between 0 and 60 s (normally) represented the ssDNA trans-cleavage rate of CRISPR/Cas12a. Data were the means and SE of three independent replicates.

## Supplementary Tables

**Supplementary Table 1.**
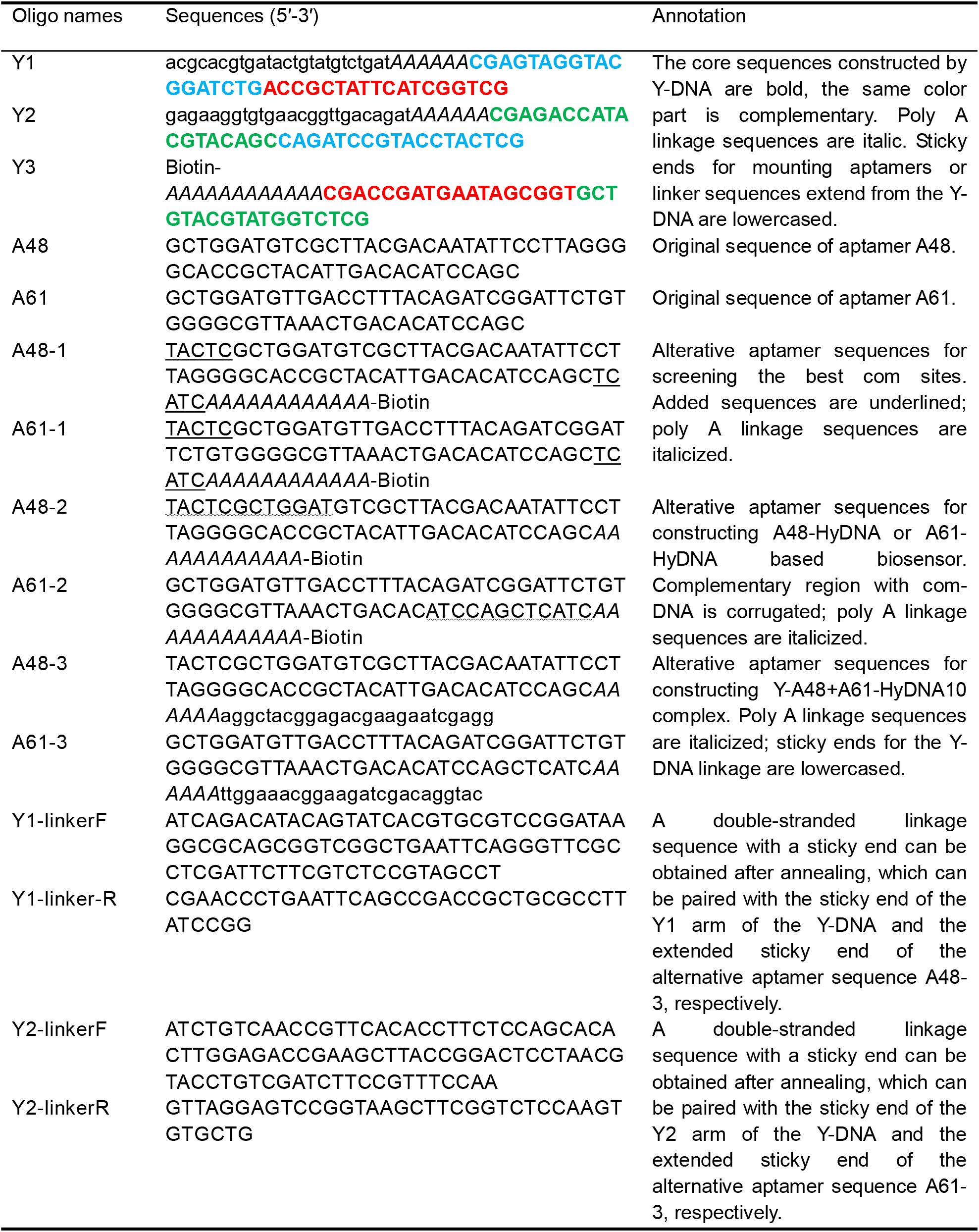
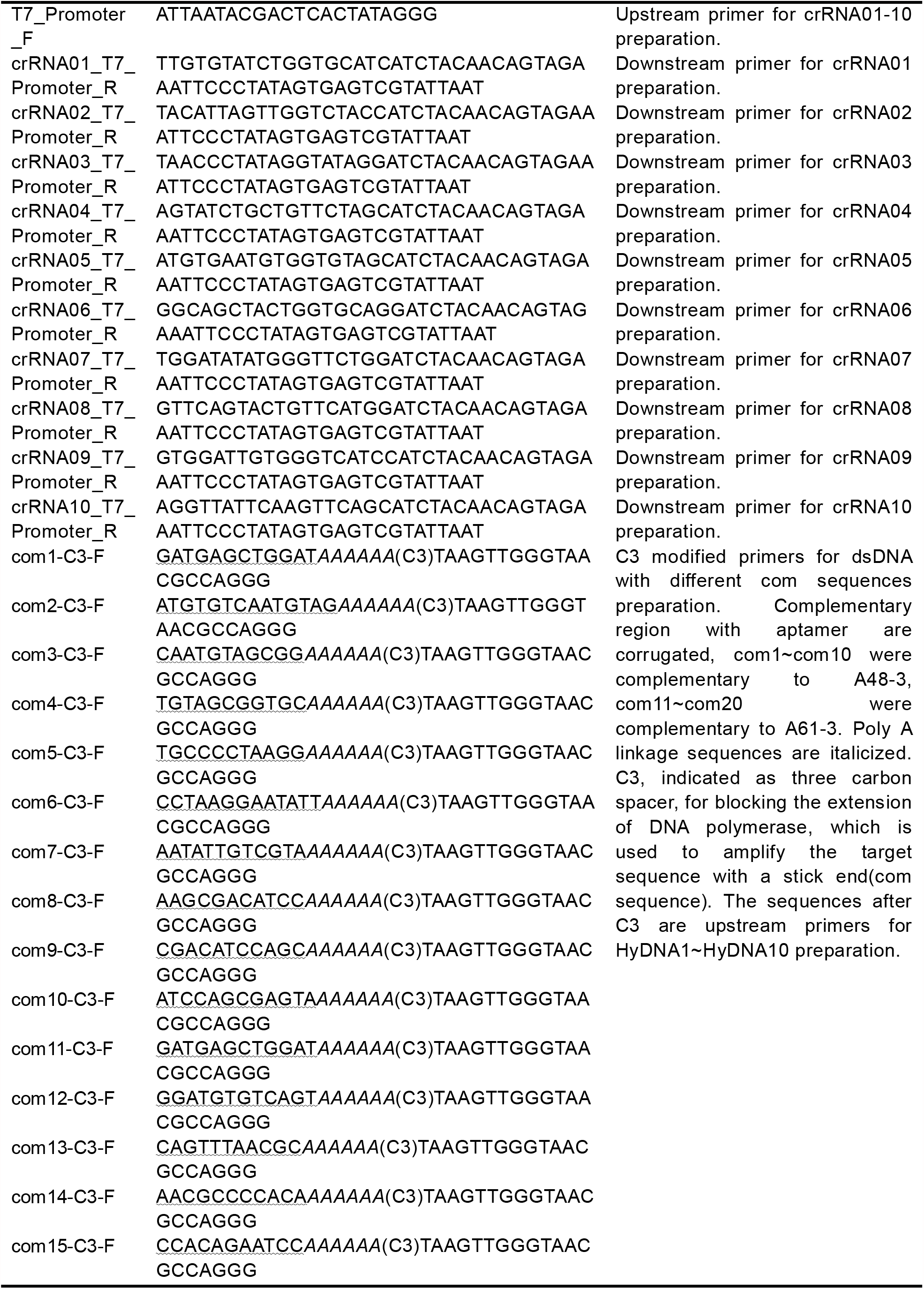
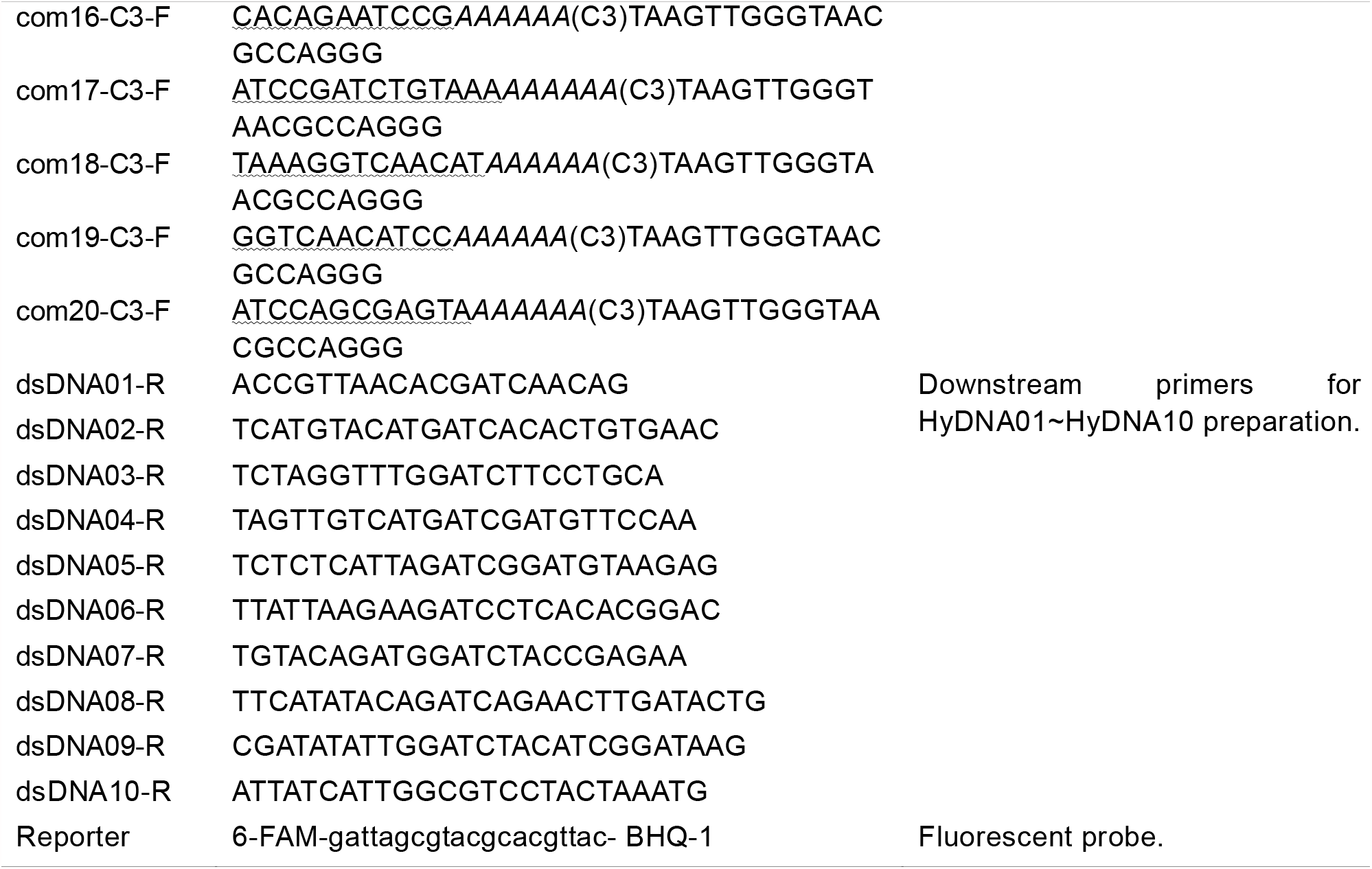
Oligonucleotides used in this study.

## Notes

### Competing Interest Statement

The authors have declared no competing interest.

### Author Declarations

Our work did not involve patient samples, and what we tested were simulated samples of inactivated viruses added to serum and saliva. The serum is purchased from Sigma-Aldrich Company Ltd. The product number is S7023. The saliva is purchased from Beijing Solarbio Science & Technology co.,ltd. The product number is A7990. The inactivated SARS-CoV-2 was prepared according to our previous study (Yang et al, Protein Cell. 2021, 12, 4.).

## References

[1] C. Wang, P. W. Horby, F. G. Hayden, G. F. Gao, The Lancet. 2020, 395, 470.

[2] J. N. Barr, R. Fearns, in Genome Stability, (Eds: I. Kovalchuk, O. Kovalchuk), Academic Press, Boston 2016, 21.

[3] J.-H. Lee, M. Choi, Y. Jung, S. K. Lee, C.-S. Lee, J. Kim, J. Kim, N. H. Kim, B.-T. Kim, H. G. Kim, Biosens. Bioelectron. 2021, 171, 112715.;

[4] a) M. M. Serrano, D. N. Rodríguez, N. T. Palop, R. O. Arenas, M. M. Córdoba, M. D. O. Mochón, C. G. Cardona, J. Clin. Virol. 2020, 129, 104529; b) J. Van Elslande, B. Decru, S. Jonckheere, E. Van Wijngaerden, E. Houben, P. Vandecandelaere, C. Indevuyst, M. Depypere, S. Desmet, E. André, M. Van Ranst, K. Lagrou, P. Vermeersch, Clin. Microbiol. Infec. 2020, 26, 1557.e1; c) Y. Boum, K. N. Fai, B. Nicolay, A. B. Mboringong, L. M. Bebell, M. Ndifon, A. Abbah, R. Essaka, L. Eteki, F. Luquero, C. Langendorf, N. F. Mbarga, R. G. Essomba, B. D. Buri, T. M. Corine, B. T. Kameni, N. Mandeng, M. Fanne, A.-C. Z.-K. Bisseck, C. B. Ndongmo, S. Eyangoh, A. Hamadou, J. P. Ouamba, M. T. Koku, R. Njouom, O. M. Claire, L. Esso, E. Epée, G. A. E. Mballa, Lancet Infect. Dis. 2021. DOI: 10.1016/S1473-3099(21)00132-8

[5] a) D. Liu, C. Ju, C. Han, R. Shi, X. Chen, D. Duan, J. Yan, X. Yan, Biosens. Bioelectron. 2021, 173, 112817; b) J. Sitjar, J.-D. Liao, H. Lee, H.-P. Tsai, J.-R. Wang, P.-Y. Liu, Biosens. Bioelectron. 2021, 181, 113153.

[6] R. L. Pinals, F. Ledesma, D. Yang, N. Navarro, S. Jeong, J. E. Pak, L. Kuo, Y.-C. Chuang, Y.-W. Cheng, H.-Y. Sun, M. P. Landry, Nano Lett. 2021, 21, 2272.

[7] T. Notomi, H. Okayama, H. Masubuchi, T. Yonekawa, K. Watanabe, N. Amino, T. Hase, Nucleic Acids Res. 2000, 28, e63.

[8] J. P. Broughton, X. Deng, G. Yu, C. L. Fasching, V. Servellita, J. Singh, X. Miao, J. A. Streithorst, A. Granados, A. Sotomayor-Gonzalez, K. Zorn, A. Gopez, E. Hsu, W. Gu, S. Miller, C.-Y. Pan, H. Guevara, D. A. Wadford, J. S. Chen, C. Y. Chiu, Nat. Biotechnol. 2020, 38, 870.

[9] J. S. Gootenberg, O. O. Abudayyeh, J. W. Lee, P. Essletzbichler, A. J. Dy, J. Joung, V. Verdine, N. Donghia, N. M. Daringer, C. A. Freije, C. Myhrvold, R. P. Bhattacharyya, J. Livny, A. Regev, E. V. Koonin, D. T. Hung, P. C. Sabeti, J. J. Collins, F. Zhang, Science. 2017, 356, 438.

[10] J. Joung, A. Ladha, M. Saito, N.-G. Kim, A. E. Woolley, M. Segel, R. P. J. Barretto, A. Ranu, R. K. Macrae, G. Faure, E. I. Ioannidi, R. N. Krajeski, R. Bruneau, M.-L. W. Huang, X. G. Yu, J. Z. Li, B. D. Walker, D. T. Hung, A. L. Greninger, K. R. Jerome, J. S. Gootenberg, O. O. Abudayyeh, F. Zhang, New Engl. J. Med. 2020, 383, 1492.

[11] X. Ding, K. Yin, Z. Li, R. V. Lalla, E. Ballesteros, M. M. Sfeir, C. Liu, Nat. Commun. 2020, 11, 4711.

[12] X. Zhao, S. Li, G. Liu, Z. Wang, Z. Yang, Q. Zhang, M. Liang, J. Liu, Z. Li, Y. Tong, G. Zhu, X. Wang, L. Jiang, W. Wang, G.-Y. Tan, L. Zhang, Sci. Bull. 2021, 66, 69.

[13] a)M. Liang, Z. Li, W. Wang, J. Liu, L. Liu, G. Zhu, L. Karthik, M. Wang, K.-F. Wang, Z. Wang, J. Yu, Y. Shuai, J. Yu, L. Zhang, Z. Yang, C. Li, Q. Zhang, T. Shi, L. Zhou, F. Xie, H. Dai, X. Liu, J. Zhang, G. Liu, Y. Zhuo, B. Zhang, C. Liu, S. Li, X. Xia, Y. Tong, Y. Liu, G. Alterovitz, G.-Y. Tan, L.-X. Zhang, Nat. Commun. 2019, 10, 3672; b)M. N. Esbin, O. N. Whitney, S. Chong, A. Maurer, X. Darzacq, R. J. R. Tjian, RNA. 2020, 26, 771.

[14] P. Fozouni, S. Son, M. Díaz de León Derby, G. J. Knott, C. N. Gray, M. V. D’Ambrosio, C. Zhao, N. A. Switz, G. R. Kumar, S. I. Stephens, D. Boehm, C.-L. Tsou, J. Shu, A. Bhuiya, M. Armstrong, A. R. Harris, P.-Y. Chen, J. M. Osterloh, A. Meyer-Franke, B. Joehnk, K. Walcott, A. Sil, C. Langelier, K. S. Pollard, E. D. Crawford, A. S. Puschnik, M. Phelps, A. Kistler, J. L. DeRisi, J. A. Doudna, D. A. Fletcher, M. Ott, Cell. 2021, 184, 323.

[15] R. Nouri, Z. Tang, M. Dong, T. Liu, A. Kshirsagar, W. Guan, Biosens. Bioelectron. 2021, 178, 113012.

[16] a) T. Hermann, D. J. Patel, Science. 2000, 287, 820; b) H. Sun, Y. Zu, Molecules. 2015, 20, 11959.

[17] Y. Xiong, J. Zhang, Z. Yang, Q. Mou, Y. Ma, Y. Xiong, Y. Lu, J. Am. Chem. Soc. 2020, 142, 207.

[18] B. Isho, K. T. Abe, M. Zuo, A. J. Jamal, B. Rathod, J. H. Wang, Z. Li, G. Chao, O. L. Rojas, Y. M. Bang, A. Pu, N. Christie-Holmes, C. Gervais, D. Ceccarelli, P. Samavarchi-Tehrani, F. Guvenc, P. Budylowski, A. Li, A. Paterson, F. Y. Yue, L. M. Marin, L. Caldwell, J. L. Wrana, K. Colwill, F. Sicheri, S. Mubareka, S. D. Gray-Owen, S. J. Drews, W. L. Siqueira, M. Barrios-Rodiles, M. Ostrowski, J. M. Rini, Y. Durocher, A. J. McGeer, J. L. Gommerman, A.-C. Gingras, Sci. Immunol. 2020, 5, eabe5511.

[19] L. Zhang, X. Fang, X. Liu, H. Ou, H. Zhang, J. Wang, Q. Li, H. Cheng, W. Zhang, Z. Luo, Chem. Commun. 2020, 56, 10235.

[20] J. Yang, P. Niu, L. Chen, L. Wang, L. Zhao, B. Huang, J. Ma, S. Hu, L. Wu, G. Wu, C. Huang, Y. Bi, W. Tan, Protein Cell. 2021, 12, 4.

[21] A. Ganguli, A. Mostafa, J. Berger, M. Y. Aydin, F. Sun, S. A. S. Ramirez, E. Valera, B. T. Cunningham, W. P. King, R. Bashir, Proc. Natl. Acad. Sci. U. S. A. 2020, 117, 22727.

[22] C. B. F. Vogels, A. F. Brito, A. L. Wyllie, J. R. Fauver, I. M. Ott, C. C. Kalinich, M. E. Petrone, A. Casanovas-Massana, M. Catherine Muenker, A. J. Moore, J. Klein, P. Lu, A. Lu-Culligan, X. Jiang, D. J. Kim, E. Kudo, T. Mao, M. Moriyama, J. E. Oh, A. Park, J. Silva, E. Song, T. Takahashi, M. Taura, M. Tokuyama, A. Venkataraman, O. E. Weizman, P. Wong, Y. Yang, N. R. Cheemarla, E. B. White, S. Lapidus, R. Earnest, B. Geng, P. Vijayakumar, C. Odio, J. Fournier, S. Bermejo, S. Farhadian, C. S. Dela Cruz, A. Iwasaki, A. I. Ko, M. L. Landry, E. F. Foxman, N. D. Grubaugh, Nat Microbiol. 2020, 5, 1299.

[23] H. Zhao, F. Liu, W. Xie, T. C. Zhou, J. OuYang, L. Jin, H. Li, C. Y. Zhao, L. Zhang, J. Wei, Y. P. Zhang, C. P. Li, Sens. Actuators. B Chem. 2021, 327, 128899.

[24] Q. Lin, D. Wen, J. Wu, L. Liu, W. Wu, X. Fang, J. Kong, Anal. Chem. 2020, 92, 9454.

[25] J. Zhong, E. L. Rosch, T. Viereck, M. Schilling, F. Ludwig, ACS Sens. 2021, 6, 976.

[26] a) H. Li, S. Xing, J. Xu, Y. He, Y. Lai, Y. Wang, G. Zhang, S. Guo, M. Deng, M. J. T. Zeng, Talanta. 2021, 221, 121670; b) H. Li, M. Li, Y. Yang, F. Wang, F. Wang, C. J. A. C. Li, Anal. Chem. 2021, 93, 3209; c) C. Niu, C. Wang, F. Li, X. Zheng, X. Xing, C. J. B. Zhang, Biosens. Bioelectron., 2021, 113196; d) B. Qiao, J. Xu, W. Yin, W. Xin, L. Ma, J. Qiao, Y. J. B. Liu, Biosens. Bioelectron. 2021, 113233.

[27] J. S. Chen, E. Ma, L. B. Harrington, M. Da Costa, X. Tian, J. M. Palefsky, J. A. Doudna, Science. 2018, 360, 436.

[28] M. Darmostuk, S. Rimpelova, H. Gbelcova, T. Ruml, Biotechnol. Adv. 2015, 33, 1141.

[29] A. J. Simon, A. Vallée-Bélisle, F. Ricci, H. M. Watkins, K. W. Plaxco, Angew. Chem. Int. Edit. 2014, 53, 9471.

[30] a)J. S. Gootenberg, O. O. Abudayyeh, M. J. Kellner, J. Joung, J. J. Collins, F. Zhang, Science 2018, 360, 439; b)Z. Ali, R. Aman, A. Mahas, G. S. Rao, M. Tehseen, T. Marsic, R. Salunke, A. K. Subudhi, S. M. Hala, S. M. Hamdan, A. Pain, F. S. Alofi, A. Alsomali, A. M. Hashem, A. Khogeer, N. A. M. Almontashiri, M. Abedalthagafi, N. Hassan, M. M. Mahfouz, Virus Res. 2020, 288, 198129

[31] H. Xu, A. Xia, D. Wang, Y. Zhang, S. Deng, W. Lu, J. Luo, Q. Zhong, F. Zhang, L. Zhou, W. Zhang, Y. Wang, C. Yang, K. Chang, W. Fu, J. Cui, M. Gan, D. Luo, M. Chen, Sci. Adv. 2020, 6, eaaz7445.

[32] Y. Li, Y. D. Tseng, S. Y. Kwon, L. d’Espaux, J. S. Bunch, P. L. McEuen, D. Luo, Nat. Mater. 2004, 3, 38.

